# The effectiveness of point of care high sensitivity troponin testing to improve Emergency Department flow: a multi-centre controlled interrupted time series

**DOI:** 10.64898/2026.08.27.26361548

**Authors:** Ryan McHenry, Andrew Saunders, Faheem Ahmad, Daniel Mackay

## Abstract

**Background:** Emergency Department (ED) crowding is an international crisis primarily driven by exit block. Point of care (POC) cardiac biomarker testing and reduced sampling intervals have been proposed to mitigate crowding by improving throughput, but whole-ED operational impacts remain poorly understood, and evaluations often rely on vulnerable observational designs. This study aimed to assess whether introducing POC high-sensitivity troponin testing and reduced sampling intervals changed whole-ED flow metrics, and to test the robustness of interrupted time series (ITS) methodology in this setting.

**Methods:** A multi-centre controlled interrupted time series (CITS) across two large urban intervention EDs and one untreated control ED in Glasgow, UK. The intervention combined whole-blood POC high-sensitivity troponin testing with a reduction in sampling intervals from 3 to 2 hours. Outcomes included daily ED admissions, mean occupancy, maximum occupancy, and mean length of stay. Analyses used a window of 120 days either side of each implementation date. Effects were evaluated using segmented ITS models, with and without controls, with permutation tests against 147 pre-intervention placebo dates. The minimum detectable effects of a similar study, applied to a national dataset, were simulated.

**Results:** Across 483,412 presentations to the intervention sites, the intervention produced no statistically significant change in any whole-ED flow metric against the untreated control at either site. Analysed alone, one intervention site appeared to show reductions in mean occupancy (−6.08, 95% CI −12.04 to −0.12) and maximum occupancy (−7.60, −14.47 to −0.73); the untreated control department produced reductions in the same direction at the same date, and both estimates attenuated to the null once the control was applied. Under a pre-specified 14-day transition specification the reductions in the untreated department reached statistical significance while those at the treated site did not. The study was limited by power due to the study window and limited control pool. Simulation demonstrated that a national dataset has the potential to provide operationally feasible and clinically important findings.

**Conclusion:** POC cardiac biomarker testing and reduced sampling intervals did not detectably improve whole-ED flow, though the design was underpowered. More importantly, uncontrolled ITS designs are highly vulnerable to confounding in complex healthcare systems; evaluations of operational interventions must utilise concurrent controls, and routinely report falsification tests.

**What is already known on this topic:**

- Emergency Department (ED) crowding is largely driven by ‘exit block’, yet throughput interventions like point of care (POC) testing are marketed to improve patient flow.
- Real-world effectiveness studies of rapid rule-out strategies for myocardial infarction have not consistently demonstrated operational improvements, particularly in systems constrained by downstream pressures.
- Single, uncontrolled interrupted time series (ITS) designs are frequently used to evaluate complex healthcare interventions despite known vulnerabilities to unmeasured confounding.

**What this study adds:**

- Implementing POC high-sensitivity troponin alongside reduced sampling intervals did not detectably improve whole-ED admissions, occupancy, or length of stay, though the study was critically underpowered to detect plausible, smaller effect sizes due to the dilution of whole-ED metrics.
- Uncontrolled analysis of one intervention site generated statistically significant apparent improvements in crowding that were not replicated in controlled analysis alongside an untreated department in the same health board, in which estimates of the same direction were obtained.
- Conventional model-based tests declared an effect in large proportions of pre-intervention dates, so falsification testing rather than model-based inference alone is required to interpret these designs.

**How this study might affect research, practice or policy:**

- Methodologically, researchers evaluating operational interventions in emergency care must be aware that uncontrolled ITS is vulnerable to confounding, consider designs using concurrent controls, and mandate the use of falsification (placebo) testing as routine practice.
- Policymakers and clinicians should consider the potential for the influence of system-wide shocks when evaluating positive results from uncontrolled, single-site before-and-after or ITS studies.
- Operational managers should recognise that throughput interventions cannot overcome whole-ED crowding driven by exit block; resources must be directed accordingly.

## Background

Emergency Department (ED) crowding is increasing internationally, and has been associated with patient harms, including excess deaths.[1,2] In this setting, crowding has mainly been attributed to output issues such as ‘exit block’, poor access to inpatient beds for those requiring admission.[2–4] However, some healthcare systems and interventions have aimed to mitigate the harms of crowding through measures that target throughput (those factors mainly affecting patient processing times), or input (those that seek to reduce presentations to emergency care).[3]

Chest pain is a common presentation to emergency care, for example, in the United States of America (USA) in 2022, chest pain comprised 5.3% of all presentations to EDs,[5] similar to other prospective studies in the United Kingdom (UK).[6,7] The requirement to rule-out myocardial infarction in this cohort is clinically and medico-legally important.[8,9] Point of care (POC) testing is an intervention that seeks to reduce time from the acquisition of a sample to the result, or improve the accessibility of such diagnostics. Therefore the use of cardiac biomarker POC testing has been investigated as an intervention with the potential to reduce ED crowding by improving throughput,[10] and manufacturers market such products as having benefits to patient flow.[11] This has prompted suggestions that the intervention may produce operationally significant reductions in ED occupancy.[12]

In clinical trials, the introduction of POC cardiac biomarker testing has been demonstrated to improve rates of successful discharge from the ED for those receiving the intervention,[13] although other studies have shown variable effects of the intervention on ED length of stay.[10,12] Likewise, reductions to sampling intervals have been shown to result in no significant improvements in ED flow metrics in systems with other, more operationally-significant, constraints.[14] It is therefore consistent with these findings that real-world effectiveness studies of various strategies to rule-out myocardial infarction have not demonstrated operational improvements across different testing platforms or test timings.[6] There are important gaps in the understanding of how the implementation of point of care cardiac biomarker testing might change the characteristics of patient flow for the whole-ED population: daily admissions, crowding and length of stay. Understanding how the implementation of such a strategy changes whole-ED characteristics is important, as it is these that are often targeted by implementing organisations.[15]

This study aimed to assess whether the introduction of point of care cardiac biomarker testing, alongside a concurrent reduction in sampling interval for patients requiring serial testing, changed whole-ED patient flow characteristics; and to assess the feasibility of applying interrupted time series methodology to the introduction of a complex intervention in the ED, including estimation of the minimum detectable effect if such a study were undertaken on a national level.

## Methods

### Setting

The intervention was instituted in two large urban EDs in Glasgow, Scotland, United Kingdom in 2025-2026. Site 1 and 2 see presentations of approximately 100,000 and 85,000 patients per year respectively. A third ED within the same health board, which did not implement point of care cardiac biomarker testing or change its sampling intervals at any point during the study period, served as an untreated control site, with approximately 57,000 presentations per year. All three EDs share the unscheduled care pathways, regional planning, and overlapping populations.

### The intervention

Prior to the intervention, the central laboratory at both intervention sites processed troponin testing using the Alinity i hs-cTnI (Abbott Laboratories, Illinois, USA) assay in plasma, while at intervention both sites implemented the same POC testing using the i- STAT Alinity hs-cTnI (Abbott Laboratories, Illinois, USA) assay in whole blood. At the point of the second intervention, both sites reduced sampling intervals for patients requiring serial tests from 3 hours to 2 hours. No other concurrent changes were made to patient pathways or clinical flow. The clinical pathways for the pre- and post-intervention period are demonstrated in Supplementary Figures 1 & 2.

The study assessed a time period from 1^st^ January 2024 to 29^th^ July 2026 inclusive, with Site 1 implementation on 12th November 2025 and Site 2 implementation on 12th March 2026. The control site was untreated throughout. Except for modelling of seasonality and the permutation tests, which include the entire study period, the analyses consider a time window of 120-days before and after the intervention. This ensures the first estimate for Site 1 reflects only the implementation of POC testing, while the estimates for Site 2 will reflect both that intervention, and the reductions in sampling intervals.

### Outcomes

As previous studies had shown reduction in admissions and this mechanism has been identified as one with the potential to ease whole-system pressures,[4,12] the primary outcome was number of admissions per day. Secondary outcomes were daily mean ED crowding (the mean number of patients in each ED on each patient arrival), maximum ED crowding (the maximum number of patients in each ED per day), and mean ED length of stay (in minutes). All outcomes were constructed at the level of the calendar day and included every attendance, not only those receiving cardiac biomarker testing.

### Statistical analyses

First, per Lopez Bernal et al.[16] a conventional single-site segmented interrupted time series was fitted separately at each intervention site, estimating the immediate step change and the change in slope at that site’s implementation date.[17] Generalised least squares models included day-of-week indicators, annual seasonality represented by Fourier terms, and a linear pre-intervention trend, with first-order autoregressive correlation structures to account for residual autocorrelation, fitted by restricted maximum likelihood.

The same model was applied to the untreated control site at each of the two implementation dates, as a negative control; any apparent effect in a department that received no intervention indicates confounding rather than treatment.[18]

Second, the analysis was repeated with a concurrent control, fitting the same segmented model to the difference between each treated site and the control site, so that any influence common to both departments was removed.[16] The two sites were treated on different dates and their effects were not assumed to be homogeneous, so site-specific terms were estimated throughout rather than a pooled intervention indicator.[19] Only the immediate step change is reported in the main text; placebo testing showed the slope terms to be the more poorly calibrated, as described below. All estimates are reported with 95% confidence intervals and p-values.

Because placebo testing showed that conventional model-based tests produce statistically significant results far more often than the nominal 5% when applied to dates on which no intervention occurred, effects were assessed by permutation tests.[20–22] Each estimated effect was compared with the distribution of effects obtained when the identical analysis was applied at three-day intervals (147 pre-intervention dates); the p-value is the proportion of these placebo analyses producing an effect at least as large as the one observed, and 95% confidence intervals contain the effect sizes that this comparison cannot distinguish from no effect.[23] Minimum detectable effects were derived from the same placebo distribution. Placebo rejection rates for every model are reported as a diagnostic of test calibration.[24] Estimates under conventional model-based inference were computed as sensitivity analyses.

Pre-specified sensitivity analyses comprised a 14-day transition period; comparison of Site 1 with the not-yet-treated Site 2, alone and combined with the control site, over the period before Site 2’s implementation; Bayesian structural time series estimation of the counterfactual using the control site as a covariate, conservative local-level trends and weakly informative spike-and-slab priors;[25] Bayesian change-point estimation;[26] and tests of parallel pre-intervention trends using permutation tests for daily slope changes in the pre-intervention period; residual autocorrelation and distributional assumptions were assessed with autocorrelation function and quantile-quantile plots of the model errors. Analyses were conducted in R (version 4.5.2) using the nlme, Rbeast and CausalImpact packages.[25–27]

### Prospective detectable effects for a national design

To assist future prospective studies, establishing the minimum detectable effects (MDEs) of an equivalent evaluation conducted across Scotland, simulations were undertaken of a projected national dataset. Annual attendances were assigned by an approximation according to routinely-available data on monthly attendances so that the network reproduces the national mixture of small rural departments and larger urban sites.[28]

For each site and outcome, daily values were generated from a site baseline scaled to annual attendance and simulated to include seasonality, day-of-week and secular trend. Residual variation was included with a shared national, and site-level, autoregressive component. Baselines, residual coefficients of variation, autoregression functions, seasonal amplitude, secular trend and cross-site correlation were calibrated from the three study EDs, by fitting the same seasonal regression used in the primary analysis to each site and outcome over the pre-intervention period.

Designs were fitted to the simulated networks using the same analysis pipeline in this study. For a single-site intervention design, a single treated site was analysed uncontrolled and against control panels of 1, 2, 5 and 10 sites, controls being those closest in annual attendance to the treated site. A staggered design was also evaluated, in which three sites spanning the size distribution were treated at 120-day intervals and each contrasted with the never-treated remainder of the network. Both designs were evaluated at analysis windows of ±120, ±240 and ±365 days.

The minimum detectable effects presented were of the same permutation-based design used for the primary analysis,[23] and are reported at 80% power, as a percentage of the simulated baseline, averaged over 15 independently simulated networks.

### Registration and approvals

The study was registered using the Open Science Framework (Registry Reference: Y5P9K). As an evaluation of an already-instituted intervention using routinely collected data, the study was defined as service evaluation and was approved by the Caldicott Guardian.[29] The study was reported in accordance with STROBE guidelines. The study was unfunded.

## Results

There were 483,412 presentations to the two intervention EDs over the time period under investigation; summary statistics of daily presentations for each site are demonstrated in Table 1 and for the control site as Supplementary Table 1.

**Table 1.**
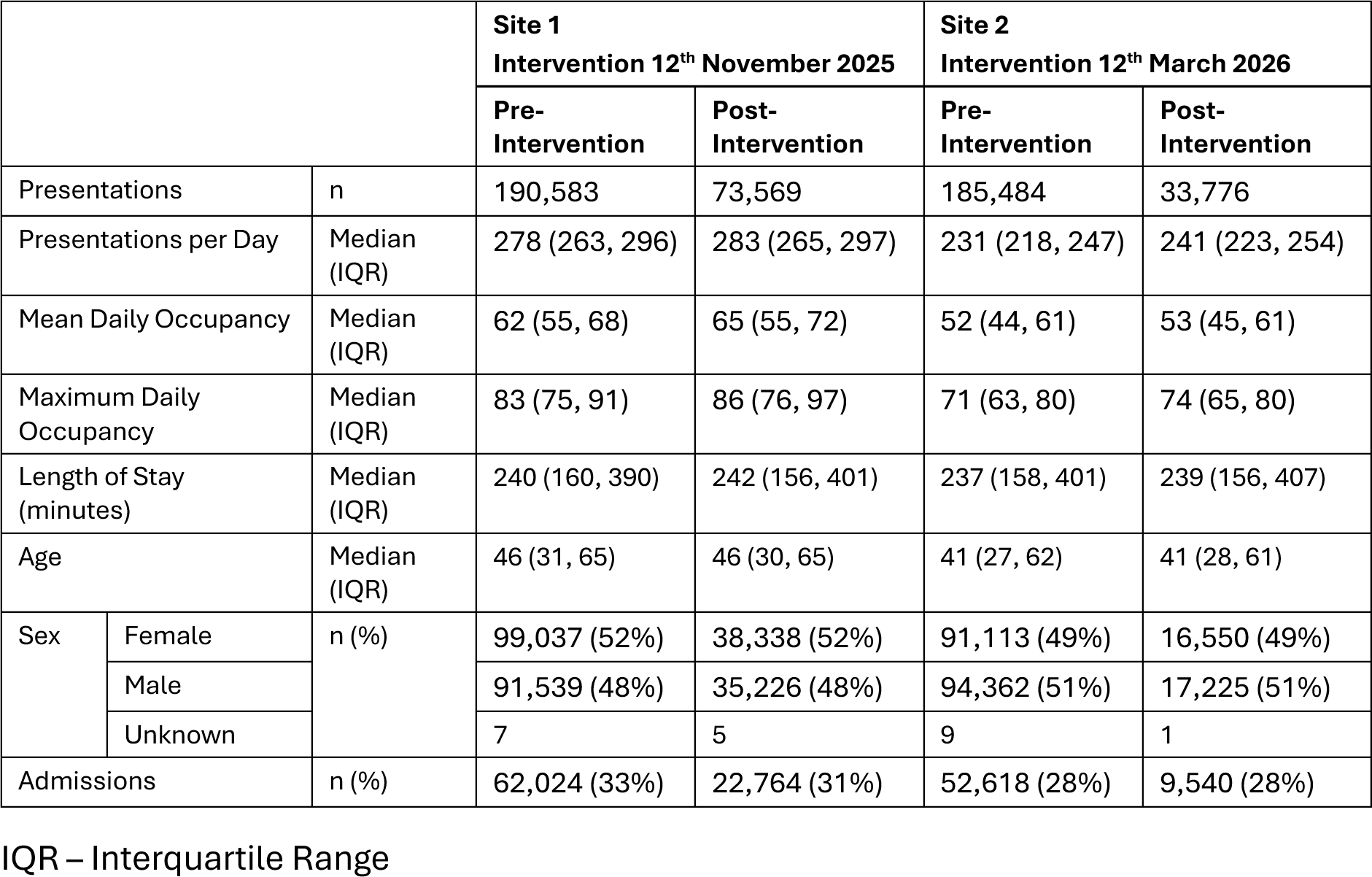
Characteristics of presentations to each intervention site from 1^st^ January 2024 to 2G^th^ July 2026, stratified by the intervention period.

Data are available on the number of troponin tests conducted through each of the pathways at Site 1 over the 75 days prior to, and following, implementation; with a mean of 42 laboratory troponin tests per day in the period prior to the intervention, and 37 POC tests per day following the intervention. This represents 14.5% and 13.3% of the total daily attendances over this time period respectively. The pathway change applied to all troponin testing. Equivalent data were not available for Site 2.

### Uncontrolled interrupted time series

Fitted at each department alone, the interrupted time series produced statistically significant reductions in mean and maximum occupancy at Site 1, and no statistically significant change in any other outcome at either site (Table 2, Figure 1). At Site 1, mean occupancy fell by 6.08 patients (95% CI −12.04 to −0.12, p=0.047) and maximum occupancy by 7.60 (95% CI −14.47 to −0.73, p=0.034). There were no significant differences in mean length of stay (-32.22 minutes; 95% CI −65.88 to 1.43, p=0.068); and daily admissions, the primary outcome, did not change (+0.99, 95% CI −5.52 to 7.49, p=0.777). Only 6 of the 147 pre-intervention placebo dates produced a reduction in mean occupancy at least as large as that observed, and only 4 produced one in maximum occupancy.

**Figure 1.**
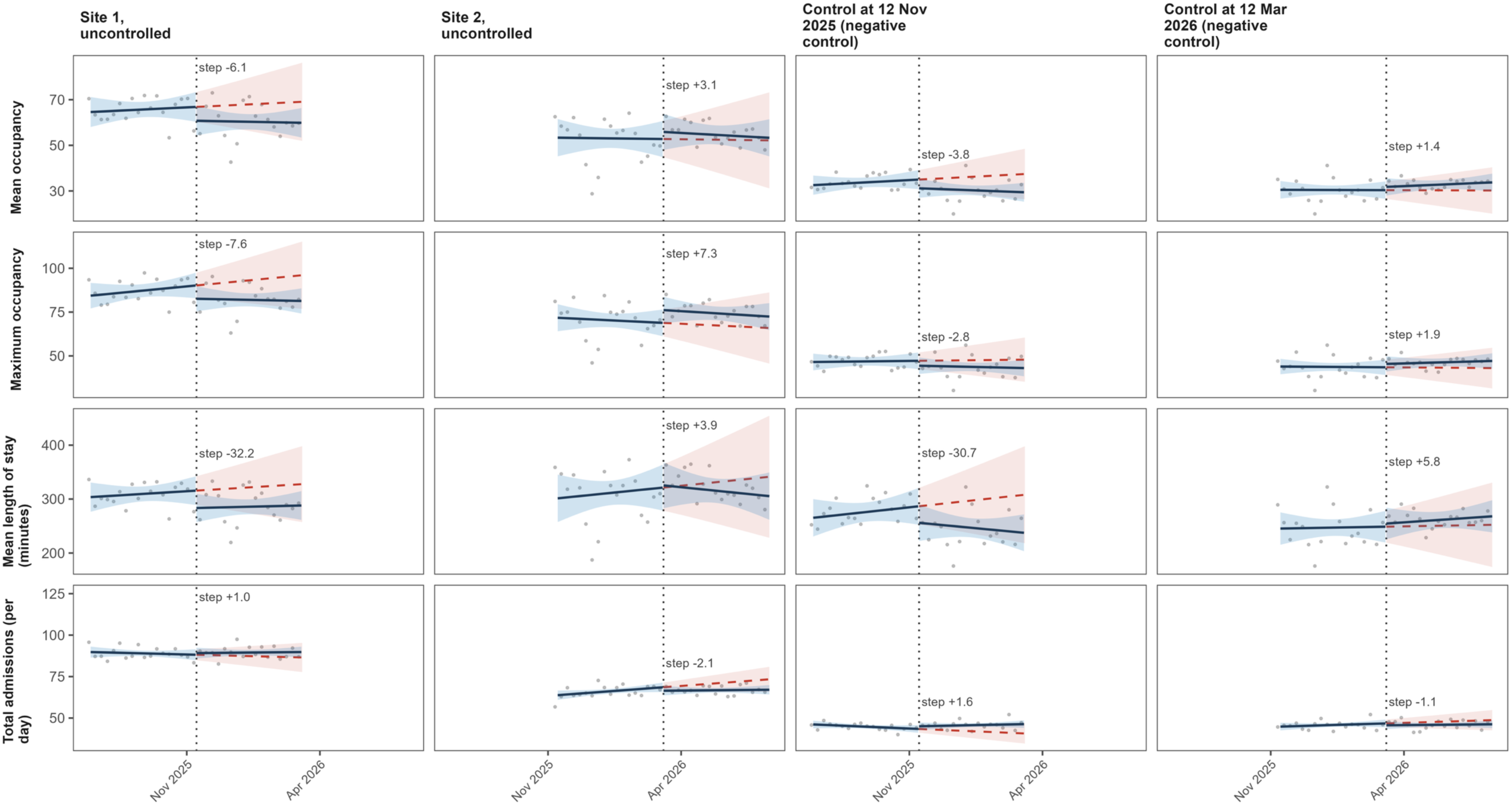
Uncontrolled interrupted time series. The two left-hand panels are the intervention sites; the two right-hand panels, labelled Control, are the untreated control site analysed with the identical model at each implementation date. Points are daily values after seasonal adjustment; solid lines are fitted pre- and post-intervention trends with G5% confidence bands and the dashed red line is the extrapolated pre-intervention trend. The dotted vertical line marks the assumed implementation date.

**Table 2.** Immediate step change in each outcome. Uncontrolled analysis, fitted separately at each intervention site and at the untreated control site at each implementation date. Controlled analysis, each intervention site compared with the untreated control. Confidence intervals and p-values are from permutation tests against 147 pre-intervention placebo dates.

| Analysis | Outcome | Series | Step (95% CI) | p |
| --- | --- | --- | --- | --- |
| Uncontrolled | Total admissions (per day) | Site 1 | 0.99 (-5.52, 7.49) | 0.777 |
|  | Total admissions (per day) | Site 2 | -2.10 (-6.23, 2.04) | 0.358 |
|  | Total admissions (per day) | Control at 12 Nov 2025 | 1.62 (-1.70, 4.94) | 0.473 |
|  | Total admissions (per day) | Control at 12 Mar 2026 | -1.15 (-4.47, 2.17) | 0.595 |
|  | Mean occupancy | Site 1 | -6.08 (-12.04, -0.12) | 0.047 |
|  | Mean occupancy | Site 2 | 3.10 (-4.88, 11.07) | 0.561 |
|  | Mean occupancy | Control at 12 Nov 2025 | -3.81 (-9.08, 1.46) | 0.142 |
|  | Mean occupancy | Control at 12 Mar 2026 | 1.44 (-3.83, 6.71) | 0.574 |
|  | Maximum occupancy | Site 1 | -7.60 (-14.47, -0.73) | 0.034 |
|  | Maximum occupancy | Site 2 | 7.30 (-0.82, 15.42) | 0.122 |
|  | Maximum occupancy | Control at 12 Nov 2025 | -2.78 (-8.33, 2.76) | 0.311 |
|  | Maximum occupancy | Control at 12 Mar 2026 | 1.93 (-3.61, 7.47) | 0.439 |
|  | Mean length of stay (minutes) | Site 1 | -32.22 (-65.88, 1.43) | 0.068 |
|  | Mean length of stay (minutes) | Site 2 | 3.94 (-43.18, 51.05) | 0.892 |
|  | Mean length of stay (minutes) | Control at 12 Nov 2025 | -30.74 (-76.17, 14.69) | 0.169 |
|  | Mean length of stay (minutes) | Control at 12 Mar 2026 | 5.79 (-39.64, 51.22) | 0.845 |
| Controlled | Total admissions (per day) | Site 1 vs Control | -0.68 (-8.38, 7.02) | 0.878 |
|  | Total admissions (per day) | Site 2 vs Control | -0.98 (-6.06, 4.09) | 0.736 |
|  | Total admissions (per day) | Site 1 vs Site 2 (to 12 Mar 2026) | 2.08 (-1.96, 6.11) | 0.392 |
|  | Total admissions (per day) | Site 1 vs Site 2+Control (to 12 Mar 2026) | 0.41 (-4.55, 5.37) | 0.905 |
|  | Mean occupancy | Site 1 vs Control | -2.00 (-9.01, 5.00) | 0.615 |
|  | Mean occupancy | Site 2 vs Control | 4.77 (-3.78, 13.32) | 0.338 |
|  | Mean occupancy | Site 1 vs Site 2 (to 12 Mar 2026) | -1.30 (-10.05, 7.44) | 0.838 |
|  | Mean occupancy | Site 1 vs Site 2+Control (to 12 Mar 2026) | -2.04 (-7.51, 3.42) | 0.615 |
|  | Maximum occupancy | Site 1 vs Control | -4.24 (-12.32, 3.84) | 0.324 |
|  | Maximum occupancy | Site 2 vs Control | 6.54 (-2.31, 15.40) | 0.176 |
|  | Maximum occupancy | Site 1 vs Site 2 (to 12 Mar 2026) | -0.96 (-9.65, 7.74) | 0.797 |
|  | Maximum occupancy | Site 1 vs Site 2+Control (to 12 Mar 2026) | -3.30 (-9.03, 2.44) | 0.284 |
|  | Mean length of stay (minutes) | Site 1 vs Control | 9.00 (-36.96, 54.96) | 0.777 |
|  | Mean length of stay (minutes) | Site 2 vs Control | 18.01 (-35.76, 71.79) | 0.574 |
|  | Mean length of stay (minutes) | Site 1 vs Site 2 (to 12 Mar 2026) | 11.98 (-39.22, 63.18) | 0.784 |
|  | Mean length of stay (minutes) | Site 1 vs Site 2+Control (to 12 Mar 2026) | 8.89 (-32.78, 50.56) | 0.689 |

At Site 2 the same model produced point estimates in the opposite direction for the crowding and length of stay outcomes, and no outcome approached statistical significance: mean occupancy +3.10 (95% CI −4.88 to 11.07, p=0.561), maximum occupancy +7.30 (95% CI −0.82 to 15.42, p=0.122), mean length of stay +3.94 minutes (95% CI −43.18 to 51.05, p=0.892), and no change in admissions (−2.10, 95% CI −6.23 to 2.04, p=0.358).

The same model was then applied to the untreated control site. At Site 1’s implementation date it produced reductions in the same direction as those estimated at Site 1 itself, of roughly half the magnitude, in a department that received no intervention: mean occupancy −3.81 (95% CI −9.08 to 1.46, p=0.142), maximum occupancy −2.78 (95% CI −8.33 to 2.76, p=0.311) and mean length of stay −30.74 minutes (95% CI −76.17 to 14.69, p=0.169). None reached statistical significance, although each did under the 14-day transition specification described below. At Site 2’s implementation date the same analysis was null for all four outcomes (Table 2).

### Controlled interrupted time series

Repeating the analysis against the untreated control left no statistically significant estimate for any outcome at either site, and removed the reductions in crowding estimated at Site 1 alone (Table 2, Figures 2 and 3). The step change in daily admissions was −0.68 per day at Site 1 (95% CI −8.38 to 7.02, p=0.878) and −0.98 at Site 2 (95% CI −6.06 to 4.09, p=0.736). Mean occupancy changed by −2.00 patients at Site 1 (95% CI −9.01 to 5.00, p=0.615) and +4.77 at Site 2 (95% CI −3.78 to 13.32, p=0.338); maximum occupancy by −4.24 (95% CI −12.32 to 3.84, p=0.324) and +6.54 (95% CI −2.31 to 15.40, p=0.176); and mean length of stay by +9.00 minutes at Site 1 (95% CI −36.96 to 54.96, p=0.777) and +18.01 minutes at Site 2 (95% CI −35.76 to 71.79, p=0.574).

**Figure 2.**
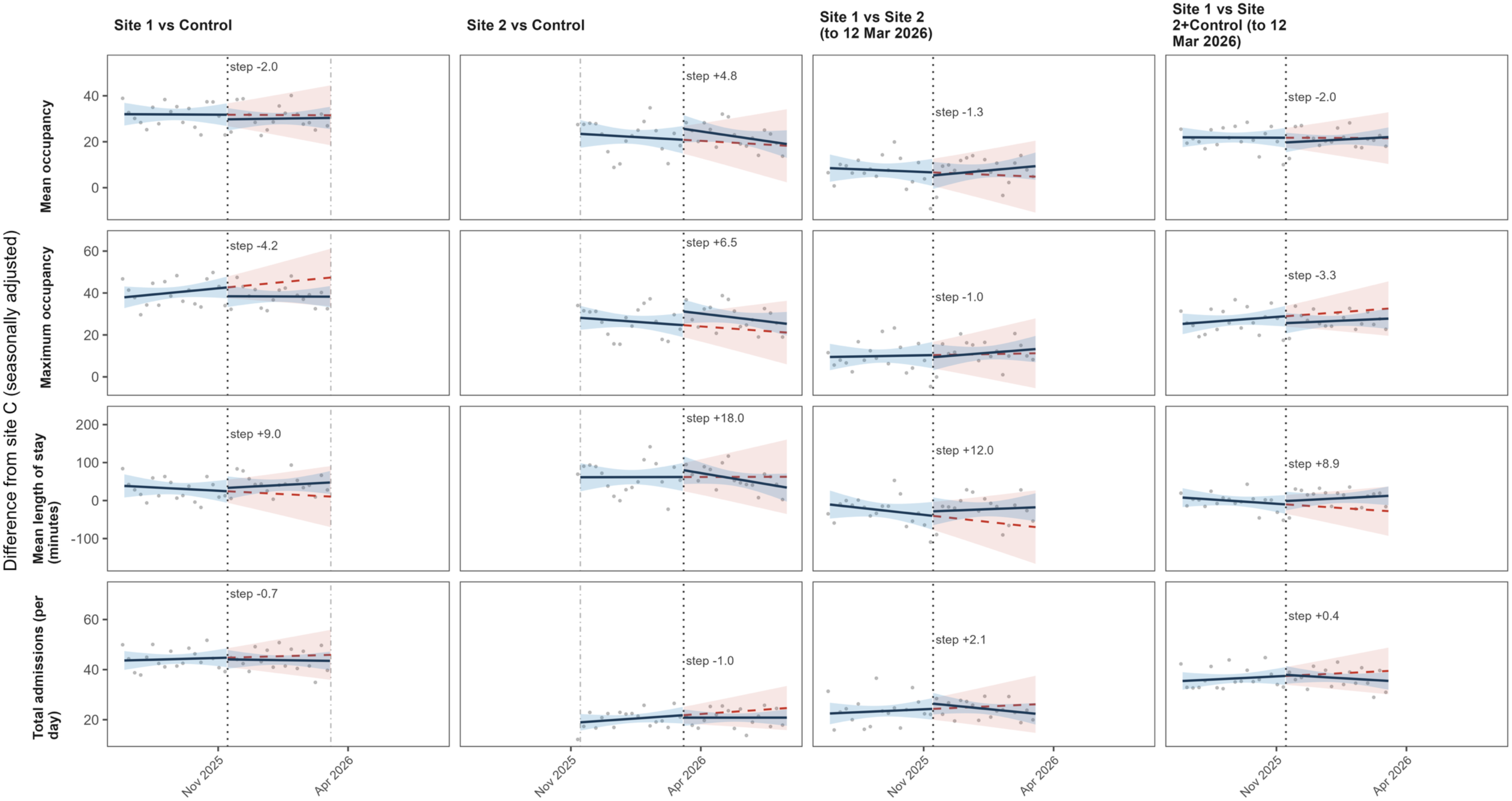
Controlled interrupted time series for each intervention site against the untreated control site, and for the additional control sets. Points are daily values after seasonal adjustment; solid lines are fitted pre- and post-intervention trends with G5% confidence bands and the dashed red line is the extrapolated pre-intervention trend. The dotted vertical line marks the assumed implementation date.

**Figure 3.**
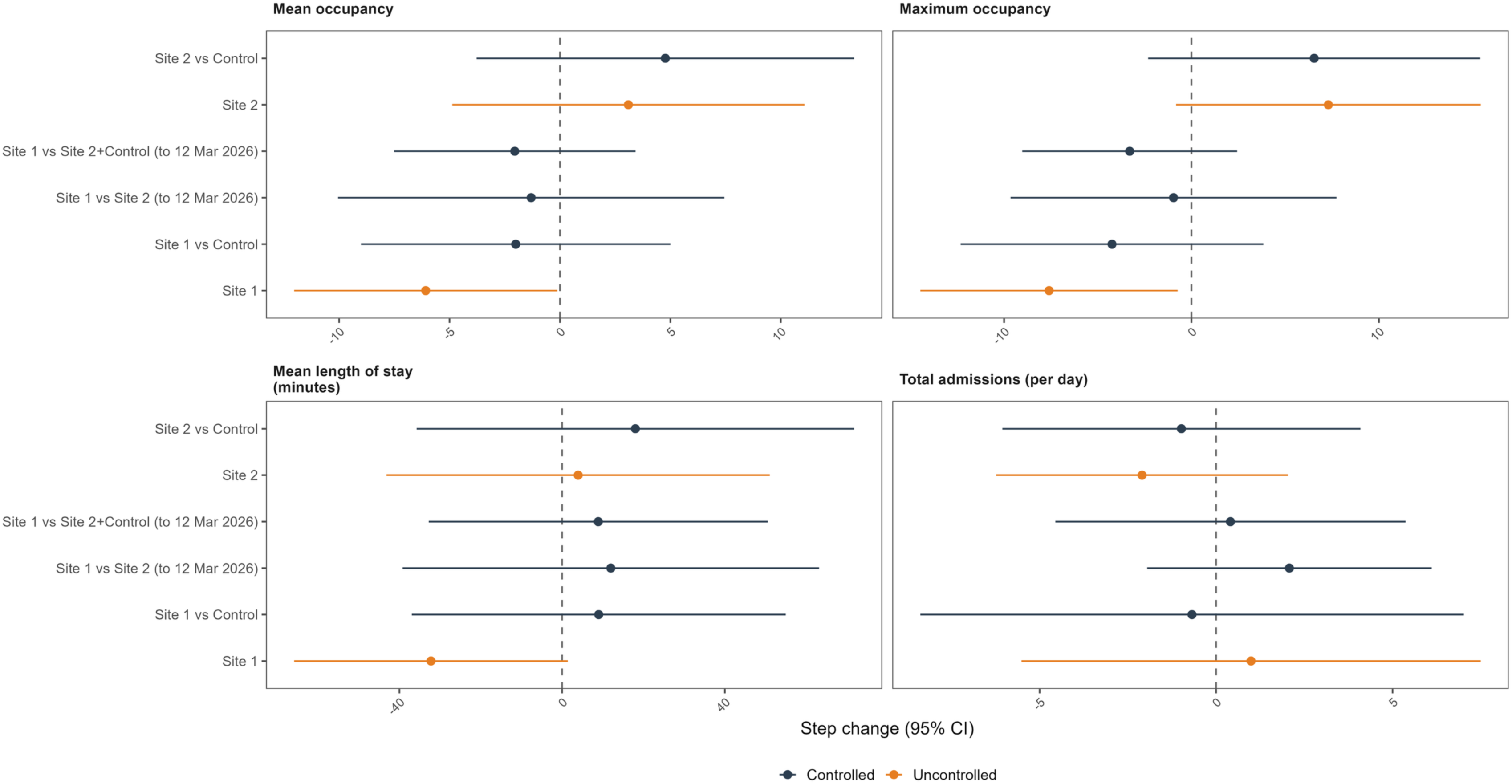
Step change estimates from the uncontrolled and controlled analyses for each outcome.

The largest controlled estimates were increases in mean length of stay of 18.01 minutes at Site 2 and 9.00 minutes at Site 1, both in the opposite direction to any intended benefit and neither statistically significant. Placebo rejection rates for the controlled step terms reached 19.1%, for daily admissions at Site 1, demonstrating misspecification of model-based inference in this setting (Supplementary Table 2).

Comparison of Site 1 with the not-yet-treated Site 2, alone and combined with the control site, produced no significant step change in any outcome (Table 2, Supplementary Table 3).

### Sensitivity analyses and power

The 14-day transition period sensitivity analysis did not change the interpretation of the controlled analysis, but it reversed which department appeared to have changed. Under that specification the reductions estimated at Site 1 alone were attenuated and no longer significant (mean occupancy −3.38, 95% CI −10.61 to 3.85, p=0.459; maximum occupancy −4.00, 95% CI −11.67 to 3.67, p=0.338), while the untreated control site returned statistically significant reductions at the same date in mean occupancy (−6.37, 95% CI −12.14 to −0.59, p=0.027) and mean length of stay (−60.66 minutes, 95% CI −109.19 to −12.13, p=0.007) (Supplementary Table 3).

Bayesian structural time series estimation using the control site as a covariate was uninformative at both sites, with wide credible intervals spanning zero on every outcome (Supplementary Table 4, Supplementary Figures 3 & 4). Bayesian change-point estimation localised structural breaks following the intervention at Site 1, but not Site 2 (Supplementary Figure 5). Other model diagnostics were acceptable (Supplementary Figures 6 & 7), and permutation tests of pre-intervention parallel trends demonstrated linearity (p>0.05) for every comparison except that for maximum occupancy at Site 1 against the control (p=0.011) (Supplementary Figure 8).

The controlled analysis had 80% power to detect step changes of 11.1 and 7.4 admissions per day at Sites 1 and 2 (12.2% and 11.2% of baseline), 10.1 and 12.4 patients in mean occupancy (16.3% and 23.6%), 11.5 and 12.8 patients in maximum occupancy (13.8% and 17.9%), and 67.2 and 78.2 minutes in mean length of stay (22.7% and 25.6%). For each intervention site versus the untreated control, every permutation-based estimate was larger than the corresponding model-based calculation, although this was not true of all the additional control sets (Supplementary Table 5).

Placebo testing of the slope terms returned statistically significant results at up to 23.1% of pre-intervention placebo dates, exceeding 10% in 15 of the 32 series and outcome combinations analysed; against a maximum of 19.1% for the step terms (Supplementary Table 2).

### Minimum detectable effects of a prospective national study

Expansion of control sites nationally was modelled. For maximum occupancy over the same ±120-day window, a single treated department analysed alone had 80% power to detect a step change of 17.4% of baseline. Adding a national control panel narrowed this; against 5 control sites the minimum detectable effect fell to 15.7% and against 10 sites to 15.0%. Widening the analysis window reduced the minimum detectable effect to a greater degree. With 10 control sites and a ±365-day window the minimum detectable effect fell to 8.3% (Supplementary Table 6). A staggered rollout to three departments at 120-day intervals, each contrasted with the never-treated remainder of the network, gave minimum detectable effects of 12.1% to 14.8% at ±120 days and 8.2% to 10.7% at ±365 days (Supplementary Table 7). The pattern was similar for the other three outcomes, and such studies are likely to be powered to detect smaller changes in admission numbers.

## Discussion

This evaluation of point of care cardiac biomarker testing, implemented alongside a reduction in sampling intervals for patients requiring serial testing, found no evidence that the intervention changed whole-ED admissions, occupancy or length of stay in two large urban EDs. No controlled estimate reached statistical significance for any of the four outcomes at either site. Analysed alone, Site 1 did produce statistically significant reductions in mean and maximum occupancy; these did not persist when the department was compared with an untreated control department in the same health board, and estimates in the same direction were obtained in that untreated department at the same date.

The untreated control site moved in the same direction as Site 1 at its implementation date despite receiving no intervention, and under the pre-specified 14-day transition specification its reductions in mean occupancy and mean length of stay reached statistical significance while those at the treated site did not. This illustrates the risk of interpreting uncontrolled interrupted time series in complex healthcare systems. That a department which received no intervention can produce the larger and more clearly significant apparent effect indicates that these estimates reflect influences common to the region rather than the intervention, and that the difference between a treated and an untreated department in the same system is more informative. While robust in certain settings, single interrupted time series designs are known to be vulnerable to this type of confounding; in one comparison against a randomised benchmark the design produced large, statistically significant estimates of the wrong sign.[30]

These findings sit alongside a trial literature in which effects have been demonstrated for the patients who receive the test rather than for the department as a whole.[13] They are consistent with real-world evaluations of rule-out strategies that have not shown operational improvement across testing platforms or test timings,[6] and with the finding that reduced sampling intervals do not improve ED flow metrics in systems constrained elsewhere.[14]

In trials of the intervention, emphasis has been placed on the importance of implementation. One described “comprehensive pre-project stakeholder discovery, engagement and collaboration” as intrinsic to the improvements demonstrated,[12] and another concluded that any improvement was “markedly dependent on its acceptance and uptake by attending personnel, and on the ED setting in which it is used”.[31] The assumption that point of care testing results in faster clinical pathways has been questioned previously in the literature,[32] and the literature on patient flow warns against expecting efficiency gains in one part of a system whose constraint lies elsewhere.[33]

### Implications for research

This study demonstrates the hazards of applying simple uncontrolled interrupted time series to complex healthcare interventions,[16] and the need to consider whether the Stable Unit Treatment Value Assumption (SUTVA) holds; that is, that a unit’s outcome does not depend on the treatments assigned to other units. This assumption can be violated where control and treated departments share a system.[34] In this study, the attenuation of the apparently significant effect for the uncontrolled analysis of Site 1 to the controlled analyses may be attributable to a whole-system shock, or that SUTVA is violated; here this would suggest that reductions in length of stay are propagated across departments, something that may be plausible in a system where ambulance bypass may be triggered.

Future research should consider an expanded panel of control sites or synthetic controls,[23] with no plausible transfer of patients between control and treated sites, as well as ensuring those studies account for within-region effects of interventions. Clinicians and policymakers should be aware of the limitations of uncontrolled interrupted time series, as there is the potential for similar magnitude effects to be seen even in sites with no intervention.

We would suggest five practical implications for evaluations of operational interventions in emergency care, which echo those in the econometric literature.

1. A panel of concurrent controls can substantially strengthen the design.[16]
2. Where these are used, both controlled and uncontrolled analyses should be reported.[16]
3. Careful consideration of study windows is required.[35]
4. Where an untreated unit exists it should be analysed as a negative control.[18]
5. The permutation distribution has the potential to improve model robustness and causal inference.[21]

### Strengths and limitations

Interrupted time series is regarded as among the strongest quasi-experimental designs where randomisation is not feasible,[17] and the staggered implementation here allowed site-specific rather than pooled estimation, avoiding the bias that arises when heterogeneous staggered effects are combined.[19] A comparable staggered approach has recently been applied to complex interventions in healthcare.[36] The principal strength of this study is the availability of an untreated department in the same system, and the pre-specification of falsification testing before any effect estimate was interpreted.

In common with many complex interventions,[37] the concurrent introduction of POC testing and a reduction in sampling intervals at Site 2 means the effect of each cannot be causally separated; this part of the evaluation represents both combined. The post-intervention period is short, so these results describe the first months of implementation rather than a settled state.

For Site 1, the controlled analysis had 80% power to detect step changes of 12.2% in daily admissions, 16.3% in mean occupancy, 13.8% in maximum occupancy and 22.7% in mean length of stay. Smaller effects cannot be excluded, and these thresholds are larger than the effects plausible for this intervention; larger studies are required to improve precision. The 120-day analysis window was chosen so that the first Site 1 estimate reflects the introduction of point of care testing alone, and this necessarily limits precision: in the national simulation, extending the window from ±120 to ±365 days reduced the minimum detectable effect for mean occupancy, and produced minimum detectable effects that are likely both operationally feasible and clinically important.

Although chest pain is likely the second-largest patient group requiring emergency care,[5] and the intervention reached a greater proportion of attendances than previous literature would predict,[7] it remains a minority of attendances, and a clinically meaningful improvement confined to that group could be diluted below detection in the whole-ED denominator. Regardless, the whole-ED scope allows assessment of unintended consequences, such as those arising from diversion of staff time to point of care testing.[32] This study does not assess the effect of the intervention for the patients receiving it, which will be considered in other work; it addresses the whole-ED effect, which is frequently the operational rationale offered for implementation.[15]

The EDs in this study, like EDs nationally, are subject to severe and persistent crowding,[2] driven by exit block, in which patients requiring admission sustain long waits for an inpatient bed.[1,38] In such a system, flow metrics are likely more amenable to interventions targeting the ‘output’ causes of poor flow.[3,33] These findings are therefore most generalisable to similar settings, and less so to departments whose crowding arises principally from poor throughput.[3]

## Conclusion

Point of care cardiac biomarker testing, including that implemented with a reduction in sampling intervals for those requiring serial testing, did not result in detectable changes to whole-ED admissions, occupancy or length of stay when compared with an untreated department in the same health board. The apparent improvements seen when one department was analysed alone were markedly attenuated when compared to a department that received no intervention.

Policymakers and operational managers should recognise that the major driver of crowding is exit block and direct resource accordingly. Those implementing POC cardiac biomarker testing to improve departmental flow should ensure their setting is configured so that a faster result can be converted into an earlier decision and departure. Evaluations of operational interventions in emergency care should use concurrent controls and report falsification tests as a matter of routine.

## Data Availability

Data are available upon reasonable request and following the appropriate ethical and governance review.

## Notes

### Competing Interest Statement

AS has received honoraria from Abbot for educational activities.
The remaining authors declare no competing interests.

### Clinical Protocols

https://osf.io/y5p9k/overview

### Author Declarations

The study was registered using the Open Science Framework (Registry Reference: Y5P9K). As an evaluation of an already-instituted intervention using routinely collected data, the study was defined as service evaluation and was approved by the Caldicott Guardian. NHS Greater Glasgow and Clyde Caldicott Guardian

