## Supplement for "The effectiveness of point of care high sensitivity troponin testing to improve Emergency Department flow: a multi-centre controlled interrupted time series"

**Supplementary Appendix.**

**Supplementary Table 1. Characteristics of presentations to the untreated control site from 1st January 2024 to 29th July 2026, stratified by the intervention period at each treated site.**

| | | | Control by Site 1 Intervention 12th November 2025 | | Control by Site 2 Intervention 12th March 2026 | |
|---|---|---|---|---|---|---|
| | | | Pre-Intervention | Post-Intervention | Pre-Intervention | Post-Intervention |
| Presentations | | n | 107,212 | 42,665 | 125,725 | 24,152 |
| Presentations per Day | | Median (IQR) | 157 (146, 170) | 162 (150, 176) | 156 (146, 169) | 173 (156, 183) |
| Mean Daily Occupancy | | Median (IQR) | 34 (29, 40) | 32 (27, 37) | 33 (28, 39) | 33 (29, 37) |
| Maximum Daily Occupancy | | Median (IQR) | 47 (41, 54) | 45 (39, 52) | 47 (40, 54) | 46 (42, 52) |
| Length of Stay (minutes) | | Median (IQR) | 232 (147, 388) | 221 (133, 348) | 231 (144, 383) | 225 (135, 350) |
| Age | | Median (IQR) | 48 (29, 68) | 47 (29, 68) | 48 (29, 68) | 46 (29, 67) |
| Sex | Female | n (%) | 55,559 (52%) | 22,564 (53%) | 65,399 (52%) | 12,724 (53%) |
| | Male | | 51,649 (48%) | 20,099 (47%) | 60,321 (48%) | 11,427 (47%) |
| | Unknown | | 4 | 2 | 5 | 1 |
| Admissions | | n (%) | 29,492 (28%) | 12,000 (28%) | 34,930 (28%) | 6,562 (27%) |

IQR – Interquartile Range

**Supplementary Table 2.** Placebo rejection rates for step and slope terms: the proportion of 147 pre-intervention placebo dates at which the conventional model-based test declared the term statistically significant at the 5% level. Rates well above 5% indicate that the conventional test is miscalibrated for that series. The two control rows are identical because they share the same permutation period, from which the placebo dates are drawn.

| Unit | Outcome_Clean | Step (immediate) | Slope (per 30 days) |
|---|---|---|---|
| Site 1, uncontrolled | Total admissions (per day) | 14.29% | 23.13% |
| Site 2, uncontrolled | Total admissions (per day) | 6.12% | 5.44% |
| Control at 12 Nov 2025 (negative control) | Total admissions (per day) | 14.29% | 21.09% |
| Control at 12 Mar 2026 (negative control) | Total admissions (per day) | 14.29% | 21.09% |
| Site 1, uncontrolled | Mean occupancy | 2.72% | 12.24% |
| Site 2, uncontrolled | Mean occupancy | 5.44% | 4.76% |
| Control at 12 Nov 2025 (negative control) | Mean occupancy | 4.76% | 13.61% |
| Control at 12 Mar 2026 (negative control) | Mean occupancy | 4.76% | 13.61% |
| Site 1, uncontrolled | Maximum occupancy | 1.36% | 11.56% |
| Site 2, uncontrolled | Maximum occupancy | 6.80% | 16.33% |
| Control at 12 Nov 2025 (negative control) | Maximum occupancy | 2.72% | 8.16% |
| Control at 12 Mar 2026 (negative control) | Maximum occupancy | 2.72% | 8.16% |
| Site 1, uncontrolled | Mean length of stay (minutes) | 5.44% | 9.52% |
| Site 2, uncontrolled | Mean length of stay (minutes) | 13.61% | 13.61% |
| Control at 12 Nov 2025 (negative control) | Mean length of stay (minutes) | 4.76% | 21.09% |
| Control at 12 Mar 2026 (negative control) | Mean length of stay (minutes) | 4.76% | 21.09% |
| Site 1 vs Control | Total admissions (per day) | 19.05% | 12.93% |
| Site 2 vs Control | Total admissions (per day) | 13.61% | 5.44% |
| Site 1 vs Site 2 (to 12 Mar 2026) | Total admissions (per day) | 0.68% | 0.00% |
| Site 1 vs Site 2+Control (to 12 Mar 2026) | Total admissions (per day) | 4.08% | 0.00% |
| Site 1 vs Control | Mean occupancy | 8.16% | 3.40% |
| Site 2 vs Control | Mean occupancy | 9.52% | 7.48% |
| Site 1 vs Site 2 (to 12 Mar 2026) | Mean occupancy | 15.65% | 2.72% |
| Site 1 vs Site 2+Control (to 12 Mar 2026) | Mean occupancy | 3.40% | 3.40% |
| Site 1 vs Control | Maximum occupancy | 6.12% | 7.48% |
| Site 2 vs Control | Maximum occupancy | 9.52% | 11.56% |
| Site 1 vs Site 2 (to 12 Mar 2026) | Maximum occupancy | 4.76% | 2.72% |
| Site 1 vs Site 2+Control (to 12 Mar 2026) | Maximum occupancy | 2.04% | 7.48% |
| Site 1 vs Control | Mean length of stay (minutes) | 10.20% | 17.01% |
| Site 2 vs Control | Mean length of stay (minutes) | 9.52% | 4.76% |
| Site 1 vs Site 2 (to 12 Mar 2026) | Mean length of stay (minutes) | 14.97% | 4.76% |
| Site 1 vs Site 2+Control (to 12 Mar 2026) | Mean length of stay (minutes) | 12.93% | 10.88% |

**Supplementary Table 3.** Full effect table: step and slope estimates for 0-day and 14-day transition periods, including the additional control sets (Site 1 versus the not-yet-treated Site 2, and versus Site 2 and the control site combined). All confidence intervals and p-values are derived from the permutation tests.

| Unit | Outcome | Term | 0-day transition: estimate (95% CI) | p-value | 14-day transition: estimate (95% CI) | p-value |
|---|---|---|---|---|---|---|
| **Uncontrolled Analysis** | | | | | | |
| Site 1, uncontrolled | Total admissions (per day) | Step (immediate) | 0.99 (-5.52, 7.49) | 0.777 | 1.12 (-6.89, 9.12) | 0.851 |
| Site 1, uncontrolled | | Slope (per 30 days) | 0.57 (-2.09, 3.23) | 0.831 | 0.63 (-2.31, 3.58) | 0.818 |
| Site 2, uncontrolled | | Step (immediate) | -2.10 (-6.23, 2.04) | 0.358 | -2.12 (-6.41, 2.17) | 0.453 |
| Site 2, uncontrolled | | Slope (per 30 days) | -1.05 (-2.73, 0.63) | 0.277 | -1.06 (-2.98, 0.86) | 0.270 |
| Control at 12 Nov 2025 (negative control) | | Step (immediate) | 1.62 (-1.70, 4.94) | 0.473 | 1.58 (-2.13, 5.30) | 0.561 |
| Control at 12 Nov 2025 (negative control) | | Slope (per 30 days) | 1.00 (-0.81, 2.82) | 0.264 | 0.95 (-0.96, 2.86) | 0.378 |
| Control at 12 Mar 2026 (negative control) | | Step (immediate) | -1.15 (-4.47, 2.17) | 0.595 | -1.15 (-4.78, 2.48) | 0.730 |
| Control at 12 Mar 2026 (negative control) | | Slope (per 30 days) | -0.33 (-2.15, 1.49) | 0.709 | -0.39 (-2.27, 1.50) | 0.696 |
| Site 1, uncontrolled | Mean occupancy | Step (immediate) | -6.08 (-12.04, -0.12) | 0.047 | -3.38 (-10.61, 3.85) | 0.459 |
| Site 1, uncontrolled | | Slope (per 30 days) | -0.78 (-4.49, 2.93) | 0.730 | -1.27 (-5.24, 2.70) | 0.608 |
| Site 2, uncontrolled | | Step (immediate) | 3.10 (-4.88, 11.07) | 0.561 | 9.02 (-0.32, 18.36) | 0.068 |
| Site 2, uncontrolled | | Slope (per 30 days) | -0.50 (-4.66, 3.67) | 0.764 | -0.90 (-5.03, 3.24) | 0.750 |
| Control at 12 Nov 2025 (negative control) | | Step (immediate) | -3.81 (-9.08, 1.46) | 0.142 | -6.37 (-12.14, -0.59) | 0.027 |
| Control at 12 Nov 2025 (negative control) | | Slope (per 30 days) | -1.05 (-4.13, 2.03) | 0.500 | -0.44 (-3.74, 2.86) | 0.750 |
| Control at 12 Mar 2026 (negative control) | | Step (immediate) | 1.44 (-3.83, 6.71) | 0.574 | 2.92 (-3.41, 9.26) | 0.338 |
| Control at 12 Mar 2026 (negative control) | | Slope (per 30 days) | 0.52 (-2.55, 3.60) | 0.736 | 0.42 (-3.00, 3.84) | 0.777 |
| Site 1, uncontrolled | Maximum occupancy | Step (immediate) | -7.60 (-14.47, -0.73) | 0.034 | -4.00 (-11.67, 3.67) | 0.338 |
| Site 1, uncontrolled | | Slope (per 30 days) | -1.77 (-5.66, 2.12) | 0.493 | -2.58 (-7.04, 1.87) | 0.385 |
| Site 2, uncontrolled | | Step (immediate) | 7.30 (-0.82, 15.42) | 0.122 | 8.60 (-1.08, 18.28) | 0.081 |
| Site 2, uncontrolled | | Slope (per 30 days) | -0.20 (-4.64, 4.25) | 0.966 | -0.40 (-5.23, 4.44) | 0.899 |
| Control at 12 Nov 2025 (negative control) | | Step (immediate) | -2.78 (-8.33, 2.76) | 0.311 | -4.63 (-10.33, 1.07) | 0.115 |
| Control at 12 Nov 2025 (negative control) | | Slope (per 30 days) | -0.51 (-3.64, 2.63) | 0.750 | -0.05 (-3.15, 3.04) | 0.973 |
| Control at 12 Mar 2026 (negative control) | | Step (immediate) | 1.93 (-3.61, 7.47) | 0.439 | 2.52 (-3.55, 8.60) | 0.419 |
| Control at 12 Mar 2026 (negative control) | | Slope (per 30 days) | 0.53 (-2.61, 3.67) | 0.736 | 0.52 (-2.78, 3.81) | 0.750 |
| Site 1, uncontrolled | Mean length of stay (minutes) | Step (immediate) | -32.22 (-65.88, 1.43) | 0.068 | -14.90 (-59.00, 29.21) | 0.493 |
| Site 1, uncontrolled | | Slope (per 30 days) | -1.84 (-18.25, 14.57) | 0.885 | -3.76 (-22.29, 14.77) | 0.818 |
| Site 2, uncontrolled | | Step (immediate) | 3.94 (-43.18, 51.05) | 0.892 | 45.41 (-14.41, 105.23) | 0.169 |
| Site 2, uncontrolled | | Slope (per 30 days) | -9.95 (-34.79, 14.89) | 0.514 | -12.55 (-37.47, 12.37) | 0.459 |
| Control at 12 Nov 2025 (negative control) | | Step (immediate) | -30.74 (-76.17, 14.69) | 0.169 | -60.66 (-109.19, -12.13) | 0.007 |
| Control at 12 Nov 2025 (negative control) | | Slope (per 30 days) | -9.99 (-34.97, 14.99) | 0.574 | -3.54 (-29.79, 22.72) | 0.797 |
| Control at 12 Mar 2026 (negative control) | | Step (immediate) | 5.79 (-39.64, 51.22) | 0.845 | 23.72 (-27.68, 75.12) | 0.446 |
| Control at 12 Mar 2026 (negative control) | | Slope (per 30 days) | 2.49 (-22.49, 27.47) | 0.946 | 0.99 (-25.81, 27.78) | 0.926 |

| | | | | | | |
|---|---|---|---|---|---|---|
| **Controlled Analysis** | | | | | | |
| Site 1 vs Control | Total admissions (per day) | Step (immediate) | -0.68 (-8.38, 7.02) | 0.878 | -0.22 (-9.43, 9.00) | 0.993 |
| Site 1 vs Control | | Slope (per 30 days) | -0.43 (-3.32, 2.46) | 0.878 | -0.46 (-3.70, 2.77) | 0.858 |
| Site 2 vs Control | | Step (immediate) | -0.98 (-6.06, 4.09) | 0.736 | -0.37 (-6.08, 5.33) | 0.885 |
| Site 2 vs Control | | Slope (per 30 days) | -0.71 (-2.53, 1.11) | 0.466 | -0.25 (-2.35, 1.84) | 0.811 |
| Site 1 vs Site 2 (to 12 Mar 2026) | | Step (immediate) | 2.08 (-1.96, 6.11) | 0.392 | 0.54 (-4.23, 5.31) | 0.872 |
| Site 1 vs Site 2 (to 12 Mar 2026) | | Slope (per 30 days) | -1.47 (-2.86, -0.08) | 0.034 | -1.12 (-3.12, 0.87) | 0.331 |
| Site 1 vs Site 2+Control (to 12 Mar 2026) | | Step (immediate) | 0.41 (-4.55, 5.37) | 0.905 | 0.03 (-5.97, 6.04) | 0.986 |
| Site 1 vs Site 2+Control (to 12 Mar 2026) | | Slope (per 30 days) | -1.11 (-2.76, 0.54) | 0.209 | -0.94 (-3.20, 1.32) | 0.358 |
| Site 1 vs Control | Mean occupancy | Step (immediate) | -2.00 (-9.01, 5.00) | 0.615 | 2.21 (-7.26, 11.68) | 0.642 |
| Site 1 vs Control | | Slope (per 30 days) | 0.22 (-2.67, 3.10) | 0.899 | -0.75 (-4.27, 2.77) | 0.709 |
| Site 2 vs Control | | Step (immediate) | 4.77 (-3.78, 13.32) | 0.338 | 5.72 (-1.87, 13.31) | 0.236 |
| Site 2 vs Control | | Slope (per 30 days) | -1.01 (-4.50, 2.48) | 0.608 | -1.67 (-4.99, 1.66) | 0.392 |
| Site 1 vs Site 2 (to 12 Mar 2026) | | Step (immediate) | -1.30 (-10.05, 7.44) | 0.838 | 4.61 (-4.80, 14.02) | 0.432 |
| Site 1 vs Site 2 (to 12 Mar 2026) | | Slope (per 30 days) | 1.48 (-1.59, 4.55) | 0.351 | 0.34 (-2.81, 3.48) | 0.845 |
| Site 1 vs Site 2+Control (to 12 Mar 2026) | | Step (immediate) | -2.04 (-7.51, 3.42) | 0.615 | 2.70 (-3.56, 8.96) | 0.500 |
| Site 1 vs Site 2+Control (to 12 Mar 2026) | | Slope (per 30 days) | 0.62 (-1.83, 3.06) | 0.568 | -0.49 (-3.48, 2.51) | 0.797 |
| Site 1 vs Control | Maximum occupancy | Step (immediate) | -4.24 (-12.32, 3.84) | 0.324 | -0.60 (-10.82, 9.62) | 0.858 |
| Site 1 vs Control | | Slope (per 30 days) | -1.23 (-4.86, 2.40) | 0.736 | -2.05 (-6.64, 2.54) | 0.493 |
| Site 2 vs Control | | Step (immediate) | 6.54 (-2.31, 15.40) | 0.176 | 5.49 (-3.24, 14.21) | 0.284 |
| Site 2 vs Control | | Slope (per 30 days) | -0.60 (-4.90, 3.71) | 0.791 | -1.49 (-5.47, 2.50) | 0.527 |
| Site 1 vs Site 2 (to 12 Mar 2026) | | Step (immediate) | -0.96 (-9.65, 7.74) | 0.797 | 5.26 (-3.98, 14.49) | 0.277 |
| Site 1 vs Site 2 (to 12 Mar 2026) | | Slope (per 30 days) | 0.75 (-3.02, 4.51) | 0.696 | -0.69 (-4.77, 3.38) | 0.770 |
| Site 1 vs Site 2+Control (to 12 Mar 2026) | | Step (immediate) | -3.30 (-9.03, 2.44) | 0.284 | 1.57 (-4.94, 8.08) | 0.642 |
| Site 1 vs Site 2+Control (to 12 Mar 2026) | | Slope (per 30 days) | -0.38 (-3.62, 2.86) | 0.824 | -1.60 (-5.38, 2.19) | 0.527 |
| Site 1 vs Control | Mean length of stay (minutes) | Step (immediate) | 9.00 (-36.96, 54.96) | 0.777 | 42.16 (-16.69, 101.00) | 0.209 |
| Site 1 vs Control | | Slope (per 30 days) | 7.05 (-20.75, 34.84) | 0.473 | -0.57 (-31.57, 30.42) | 0.973 |
| Site 2 vs Control | | Step (immediate) | 18.01 (-35.76, 71.79) | 0.574 | 21.56 (-23.40, 66.52) | 0.581 |
| Site 2 vs Control | | Slope (per 30 days) | -11.59 (-35.20, 12.01) | 0.419 | -15.77 (-38.34, 6.80) | 0.182 |
| Site 1 vs Site 2 (to 12 Mar 2026) | | Step (immediate) | 11.98 (-39.22, 63.18) | 0.784 | 48.91 (-7.47, 105.29) | 0.101 |
| Site 1 vs Site 2 (to 12 Mar 2026) | | Slope (per 30 days) | 10.12 (-7.76, 28.00) | 0.297 | 2.27 (-15.94, 20.47) | 0.838 |
| Site 1 vs Site 2+Control (to 12 Mar 2026) | | Step (immediate) | 8.89 (-32.78, 50.56) | 0.689 | 42.41 (-4.58, 89.39) | 0.095 |
| Site 1 vs Site 2+Control (to 12 Mar 2026) | | Slope (per 30 days) | 7.92 (-10.69, 26.53) | 0.493 | 0.10 (-20.13, 20.33) | 0.993 |

**Supplementary Table 4.** Bayesian structural time series summary. Credible intervals are wide on every outcome and the analysis is uninformative rather than supportive of the null.

| Series | Outcome | Observed | Predicted | Absolute effect (95% CrI) |
|---|---|---|---|---|
| Site 1 vs Control | Admissions | 88.66 | 84.29 | 4.37 (−16.76 to 26.04) |
| Site 1 vs Control | Mean occupancy | 60.46 | 69.42 | −8.96 (−77.99 to 58.99) |
| Site 1 vs Control | Maximum occupancy | 81.25 | 91.59 | −10.34 (−73.37 to 53.91) |
| Site 1 vs Control | Length of stay | 299.2 | 323.8 | −24.6 (−321.6 to 283.2) |
| Site 2 vs Control | Admissions | 68.03 | 68.08 | −0.05 (−18.30 to 18.07) |
| Site 2 vs Control | Mean occupancy | 53.57 | 56.98 | −3.41 (−71.52 to 63.82) |
| Site 2 vs Control | Maximum occupancy | 73.43 | 75.02 | −1.60 (−53.98 to 50.67) |
| Site 2 vs Control | Length of stay | 303.4 | 329.3 | −25.8 (−383.8 to 350.3) |
| Control at 12 Nov 2025 | Admissions | 45.33 | 44.56 | 0.76 (−10.98 to 12.32) |
| Control at 12 Nov 2025 | Mean occupancy | 30.66 | 32.97 | −2.31 (−32.02 to 26.79) |
| Control at 12 Nov 2025 | Maximum occupancy | 43.82 | 45.37 | −1.55 (−29.50 to 26.22) |
| Control at 12 Nov 2025 | Length of stay | 261.1 | 259.2 | 1.9 (−283.7 to 293.3) |

CrI – Credible Interval

**Supplementary Table 5.** Minimum detectable effects at 80% power for the step change in each outcome, derived from the permutation tests, and from the model alone.

| | | | Permutation-based | | Model-based | |
|---|---|---|---|---|---|---|
| Series | Outcome | Baseline level | MDE | % of baseline | MDE | % of baseline |
| **Uncontrolled Analysis** | | | | | | |
| Site 1, uncontrolled | Total admissions (per day) | 91.2 | 9.3 | 10.2% | 6.7 | 7.3% |
| Site 2, uncontrolled | Total admissions (per day) | 65.7 | 5.9 | 9.0% | 5.8 | 8.8% |
| Control at 12 Nov 2025 (negative control) | Total admissions (per day) | 43.4 | 4.9 | 11.2% | 4.6 | 10.7% |
| Control at 12 Mar 2026 (negative control) | Total admissions (per day) | 43.7 | 4.9 | 11.1% | 4.7 | 10.7% |
| Site 1, uncontrolled | Mean occupancy | 61.7 | 8.8 | 14.3% | 12.4 | 20.1% |
| Site 2, uncontrolled | Mean occupancy | 52.6 | 11.5 | 21.8% | 14.8 | 28.2% |
| Control at 12 Nov 2025 (negative control) | Mean occupancy | 34.6 | 7.5 | 21.7% | 8.1 | 23.5% |
| Control at 12 Mar 2026 (negative control) | Mean occupancy | 34.0 | 7.5 | 22.1% | 7.5 | 22.0% |
| Site 1, uncontrolled | Maximum occupancy | 83.3 | 9.8 | 11.8% | 14.2 | 17.1% |
| Site 2, uncontrolled | Maximum occupancy | 71.5 | 11.8 | 16.5% | 15.0 | 21.0% |
| Control at 12 Nov 2025 (negative control) | Maximum occupancy | 48.0 | 7.9 | 16.4% | 9.4 | 19.5% |
| Control at 12 Mar 2026 (negative control) | Maximum occupancy | 47.4 | 7.9 | 16.7% | 8.7 | 18.3% |
| Site 1, uncontrolled | Mean length of stay (minutes) | 295.8 | 48.5 | 16.4% | 50.8 | 17.2% |
| Site 2, uncontrolled | Mean length of stay (minutes) | 305.2 | 69.3 | 22.7% | 79.0 | 25.9% |
| Control at 12 Nov 2025 (negative control) | Mean length of stay (minutes) | 289.7 | 64.2 | 22.2% | 64.1 | 22.1% |
| Control at 12 Mar 2026 (negative control) | Mean length of stay (minutes) | 285.4 | 64.2 | 22.5% | 56.6 | 19.8% |
| **Controlled Analysis** | | | | | | |
| Site 1 vs Control | Total admissions (per day) | 91.2 | 11.1 | 12.2% | 7.6 | 8.4% |
| Site 2 vs Control | Total admissions (per day) | 65.7 | 7.4 | 11.2% | 6.7 | 10.2% |
| Site 1 vs Site 2 (to 12 Mar 2026) | Total admissions (per day) | 91.2 | 5.8 | 6.4% | 8.8 | 9.6% |
| Site 1 vs Site 2+Control (to 12 Mar 2026) | Total admissions (per day) | 91.2 | 7.2 | 7.9% | 7.2 | 7.8% |
| Site 1 vs Control | Mean occupancy | 61.7 | 10.1 | 16.3% | 9.8 | 15.9% |
| Site 2 vs Control | Mean occupancy | 52.6 | 12.4 | 23.6% | 11.8 | 22.5% |
| Site 1 vs Site 2 (to 12 Mar 2026) | Mean occupancy | 61.7 | 12.8 | 20.7% | 11.8 | 19.2% |
| Site 1 vs Site 2+Control (to 12 Mar 2026) | Mean occupancy | 61.7 | 8.1 | 13.1% | 8.5 | 13.8% |
| Site 1 vs Control | Maximum occupancy | 83.3 | 11.5 | 13.8% | 10.4 | 12.5% |
| Site 2 vs Control | Maximum occupancy | 71.5 | 12.8 | 17.9% | 11.5 | 16.1% |
| Site 1 vs Site 2 (to 12 Mar 2026) | Maximum occupancy | 83.3 | 12.4 | 14.9% | 12.7 | 15.2% |
| Site 1 vs Site 2+Control (to 12 Mar 2026) | Maximum occupancy | 83.3 | 8.3 | 10.0% | 9.9 | 11.9% |
| Site 1 vs Control | Mean length of stay (minutes) | 295.8 | 67.2 | 22.7% | 59.8 | 20.2% |
| Site 2 vs Control | Mean length of stay (minutes) | 305.2 | 78.2 | 25.6% | 72.7 | 23.8% |
| Site 1 vs Site 2 (to 12 Mar 2026) | Mean length of stay (minutes) | 295.8 | 74.9 | 25.3% | 72.0 | 24.3% |
| Site 1 vs Site 2+Control (to 12 Mar 2026) | Mean length of stay (minutes) | 295.8 | 59.9 | 20.2% | 48.9 | 16.5% |

MDE – Minimum Detectable Effect

**Supplementary Table 6. Minimum detectable effects of a prospective national study assessing the impact of an intervention conducted at a single site, stratified by number of controls and time-window under investigation. Results derived from a simulation of a national Scottish study.**

| | | Change in baseline for each outcome (%) | | |
|---|---|---|---|---|
| Number of Controls | Outcome | +/-120 days | +/-240 days | +/-365 days |
| 0 | Maximum Occupancy | 17.44% | 12.70% | 10.20% |
| 1 | Maximum Occupancy | 20.27% | 14.28% | 11.10% |
| 2 | Maximum Occupancy | 17.69% | 12.52% | 9.78% |
| 5 | Maximum Occupancy | 15.74% | 10.95% | 8.59% |
| 10 | Maximum Occupancy | 15.03% | 10.54% | 8.29% |
| 0 | Mean Occupancy | 21.34% | 16.26% | 12.88% |
| 1 | Mean Occupancy | 24.48% | 17.86% | 14.20% |
| 2 | Mean Occupancy | 21.68% | 15.67% | 12.38% |
| 5 | Mean Occupancy | 18.82% | 13.68% | 10.83% |
| 10 | Mean Occupancy | 18.04% | 13.02% | 10.39% |
| 0 | Mean Length of Stay (minutes) | 21.03% | 15.90% | 12.49% |
| 1 | Mean Length of Stay (minutes) | 26.98% | 20.12% | 15.79% |
| 2 | Mean Length of Stay (minutes) | 23.55% | 17.41% | 13.72% |
| 5 | Mean Length of Stay (minutes) | 21.51% | 15.72% | 12.26% |
| 10 | Mean Length of Stay (minutes) | 20.65% | 15.00% | 11.64% |
| 0 | Total Admissions (per day) | 7.95% | 5.59% | 4.42% |
| 1 | Total Admissions (per day) | 10.16% | 7.24% | 5.78% |
| 2 | Total Admissions (per day) | 9.13% | 6.47% | 5.14% |
| 5 | Total Admissions (per day) | 8.09% | 5.67% | 4.49% |
| 10 | Total Admissions (per day) | 7.89% | 5.53% | 4.35% |

**Supplementary Table 7. Minimum detectable effects of a prospective national study assessing the impact of an intervention staggered over 3 sites at 120-day intervals, with national controls, stratified by time-window under investigation. Results derived from a simulation of a national Scottish study.**

| Outcome | Site | +/-120 days | +/-240 days | +/-365 days |
|---|---|---|---|---|
| Maximum Occupancy | Site 1 | 13.55% | 11.27% | 10.70% |
| Maximum Occupancy | Site 2 | 12.05% | 9.21% | 8.41% |
| Maximum Occupancy | Site 3 | 14.84% | 10.40% | 8.20% |
| Mean Occupancy | Site 1 | 17.63% | 15.08% | 14.45% |
| Mean Occupancy | Site 2 | 16.23% | 12.47% | 11.22% |
| Mean Occupancy | Site 3 | 17.57% | 12.74% | 10.15% |
| Mean Length of Stay (minutes) | Site 1 | 20.27% | 16.47% | 15.82% |
| Mean Length of Stay (minutes) | Site 2 | 18.04% | 14.17% | 12.80% |
| Mean Length of Stay (minutes) | Site 3 | 19.60% | 14.31% | 11.11% |
| Total Admissions (per day) | Site 1 | 11.24% | 9.50% | 9.05% |
| Total Admissions (per day) | Site 2 | 9.23% | 6.63% | 6.06% |
| Total Admissions (per day) | Site 3 | 7.71% | 5.37% | 4.22% |

**Supplementary Figure 1.** Clinical pathway prior to the intervention.

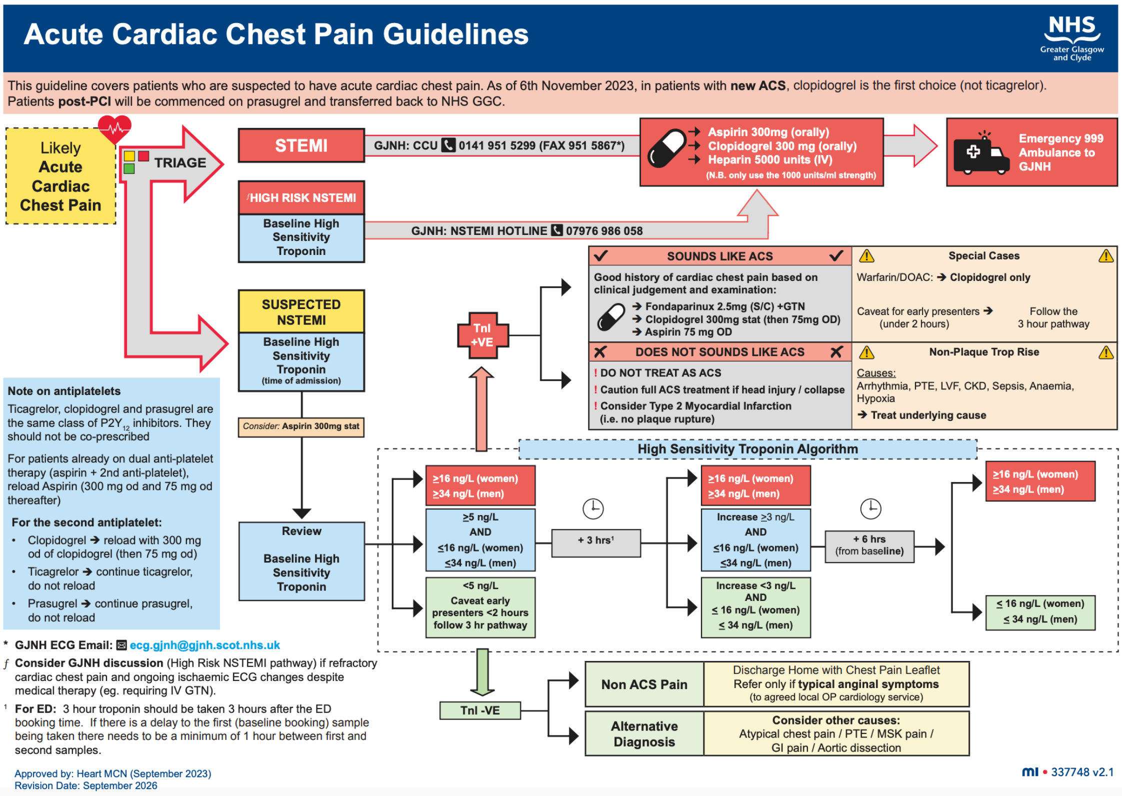


**Supplementary Figure 2.** Clinical pathway following the March 12th intervention. The only amendment from that of November 12th was of the sampling interval.

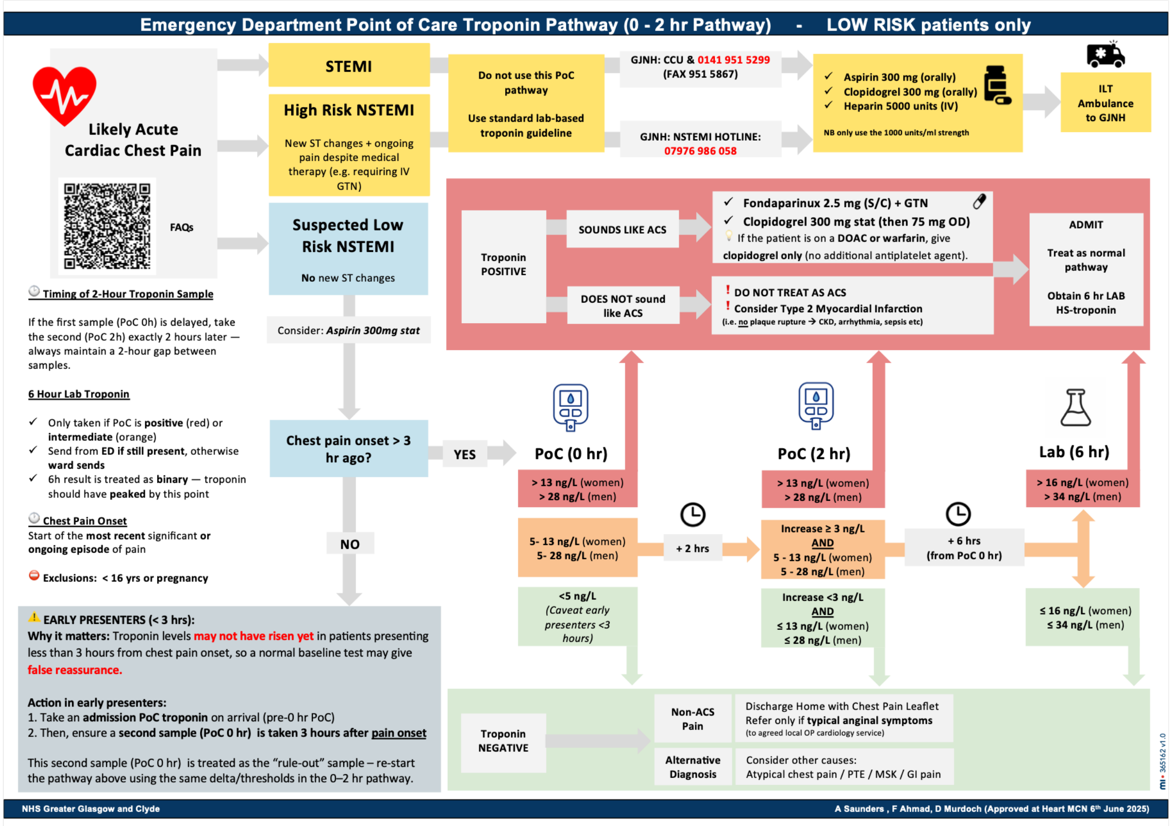

**Supplementary Figure 3.** Bayesian structural time series pointwise effect estimates against the control-site counterfactual. Shaded area is 95% credible band.

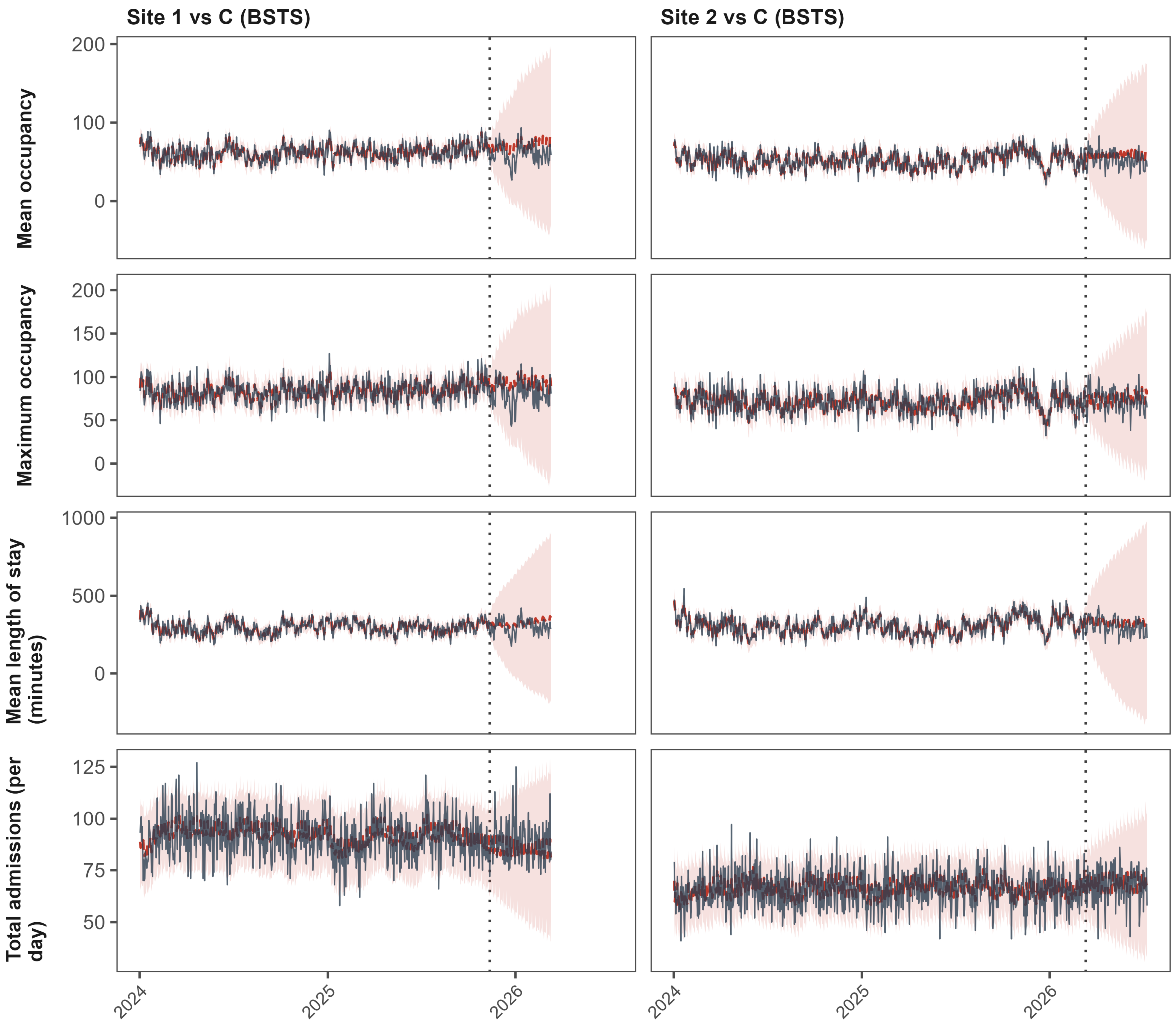

**Supplementary Figure 4.** Bayesian structural time series cumulative effect estimates against the control-site counterfactual.

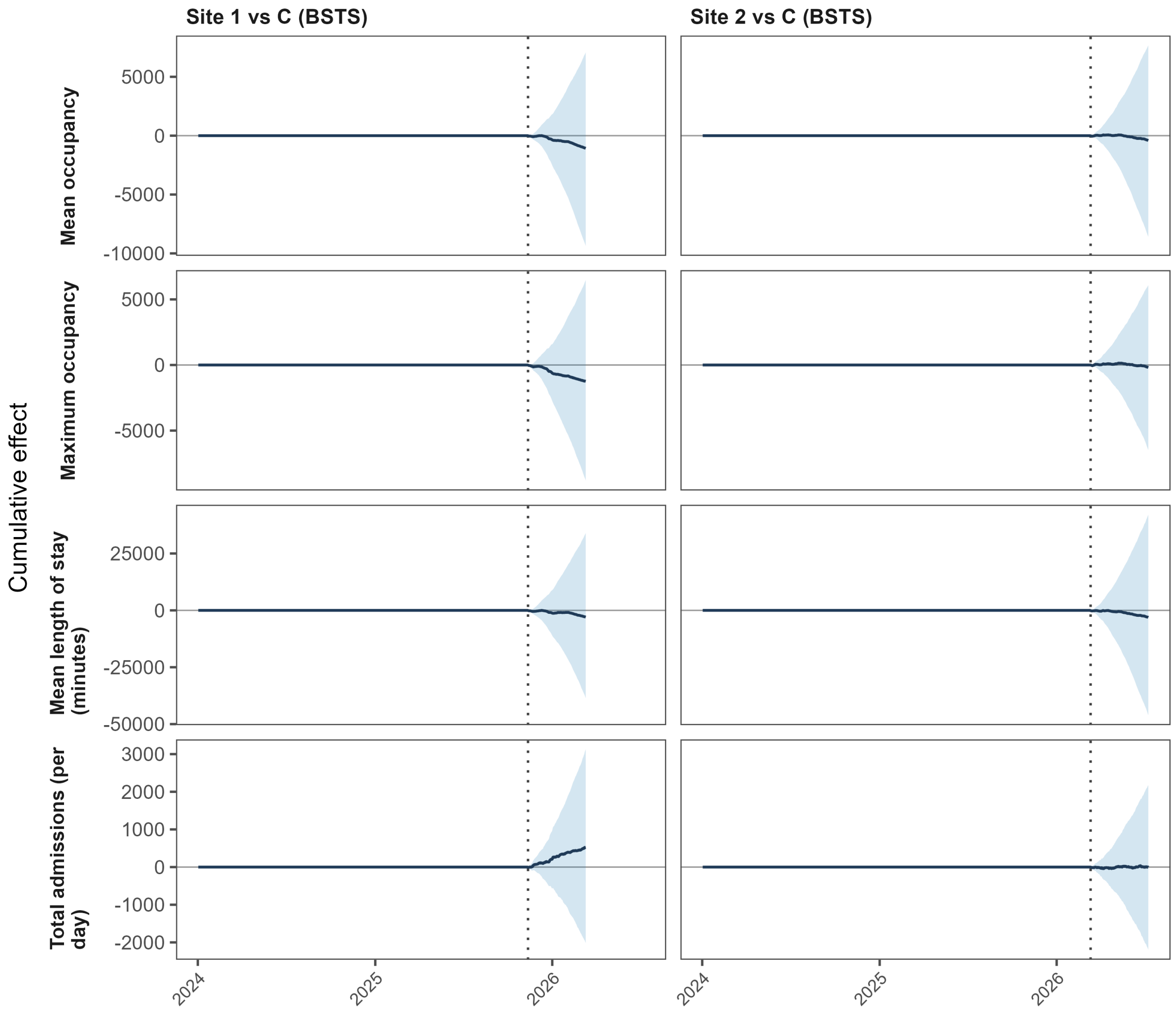

**Supplementary Figure 5.** Bayesian change-point estimates for each site and outcome. The shaded band indicates +-14 days around the actual implementation date.

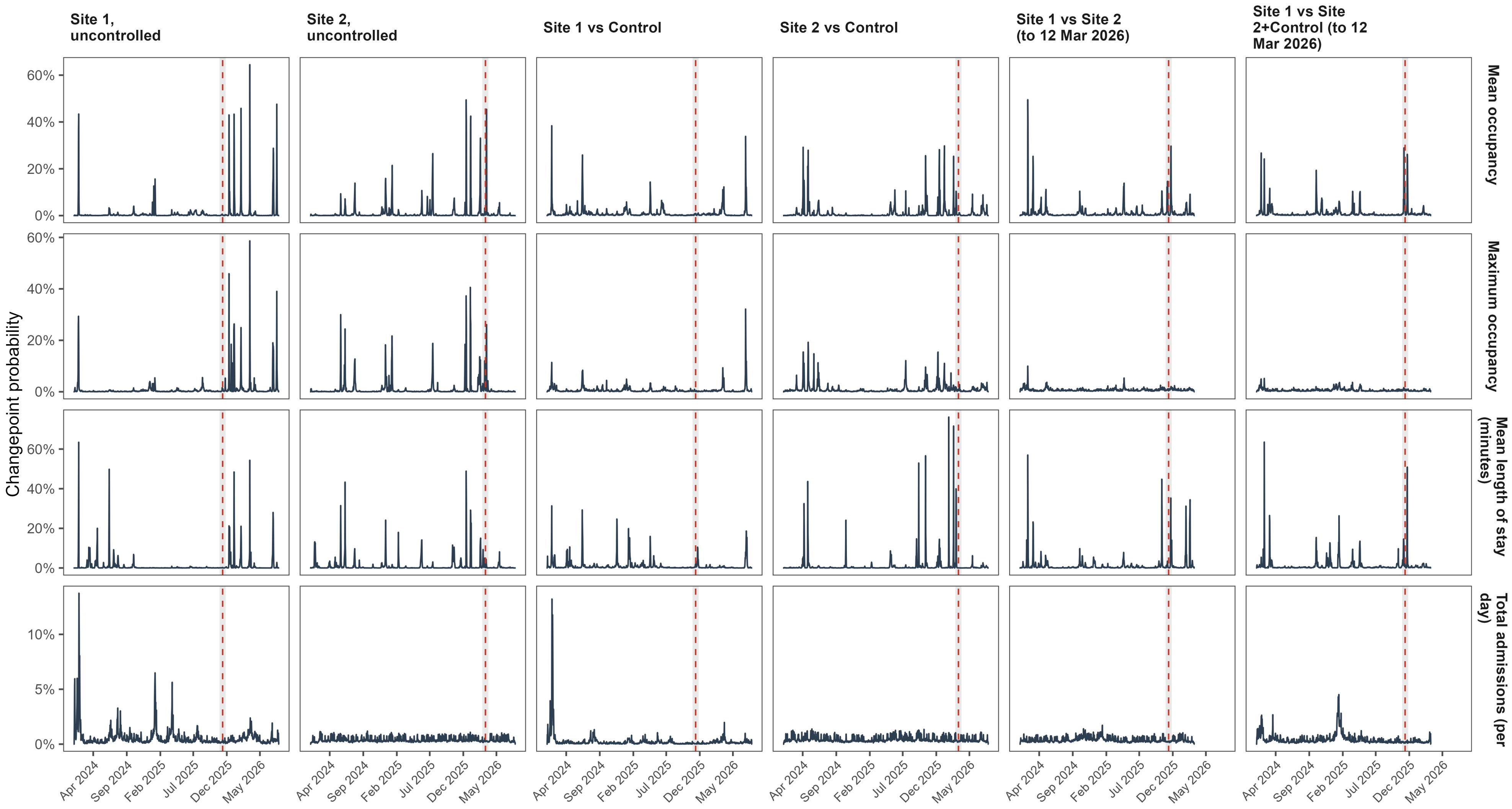

**Supplementary Figure 6.** Autocorrelation function plots of model residuals for every unit and outcome.

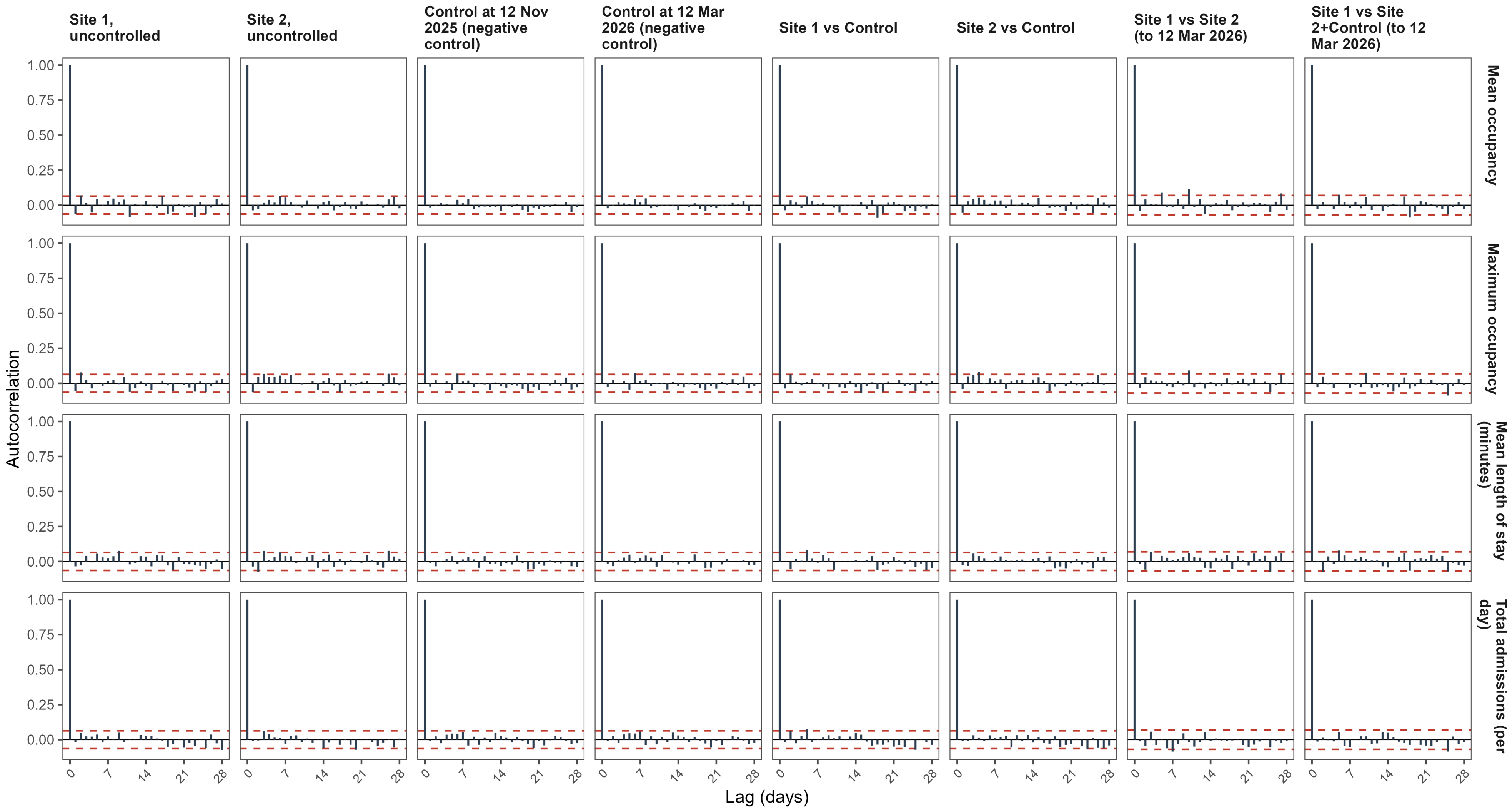

**Supplementary Figure 7.** Quantile-quantile plots of model residuals for every unit and outcome.

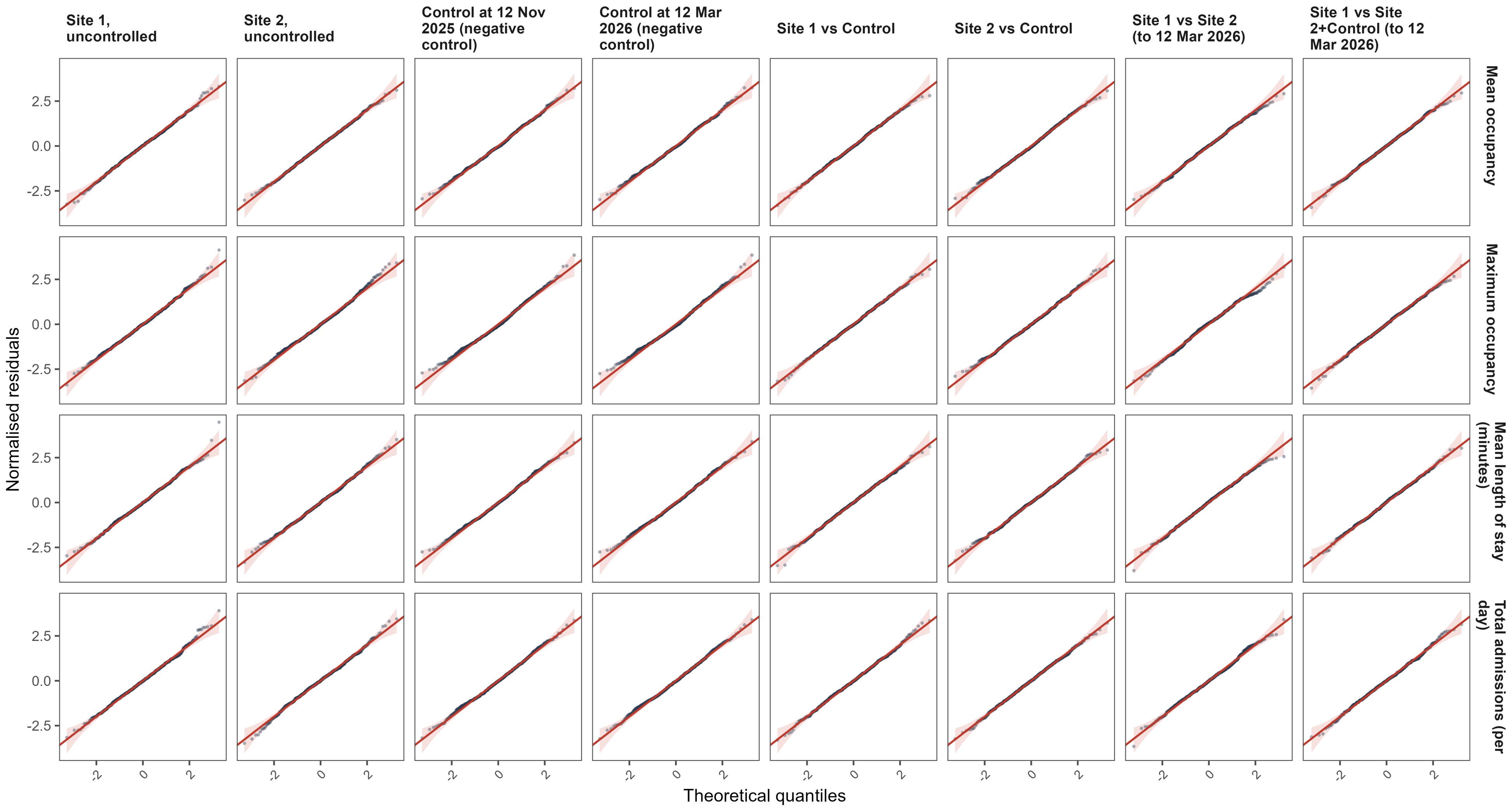

**Supplementary Figure 8.** Plotted parallel trends for each modelled comparison and outcome. Permutation tests demonstrate no significant diverging trends for any outcome-comparison pair except for maximum occupancy in Site 1 against Control. Points are the weekly mean of aggregated daily slopes for legibility.

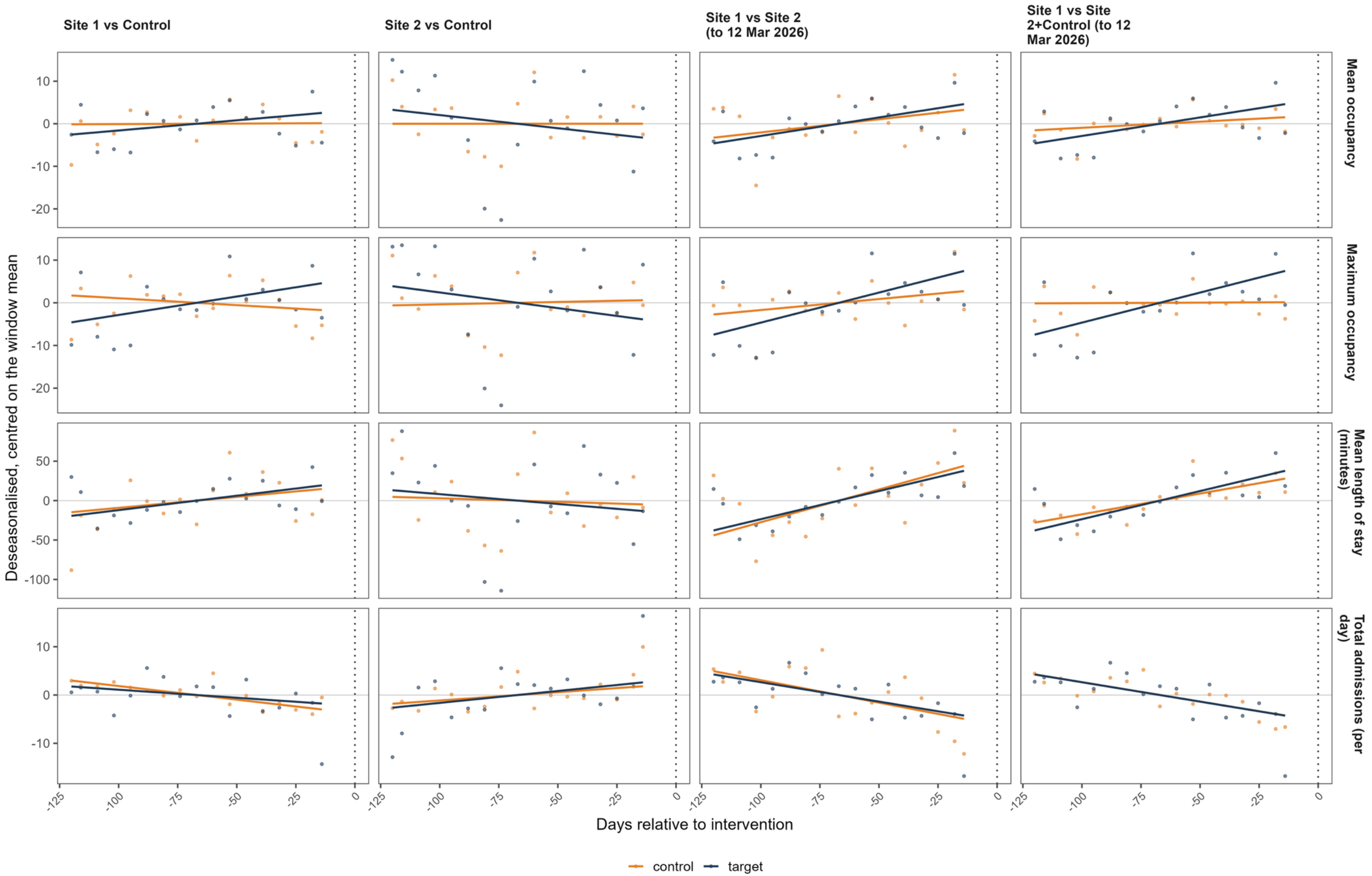

**Supplementary Figure 9.** Placebo sweep: step and slope estimates obtained at each of 147 pre-intervention placebo dates, for every unit and outcome.

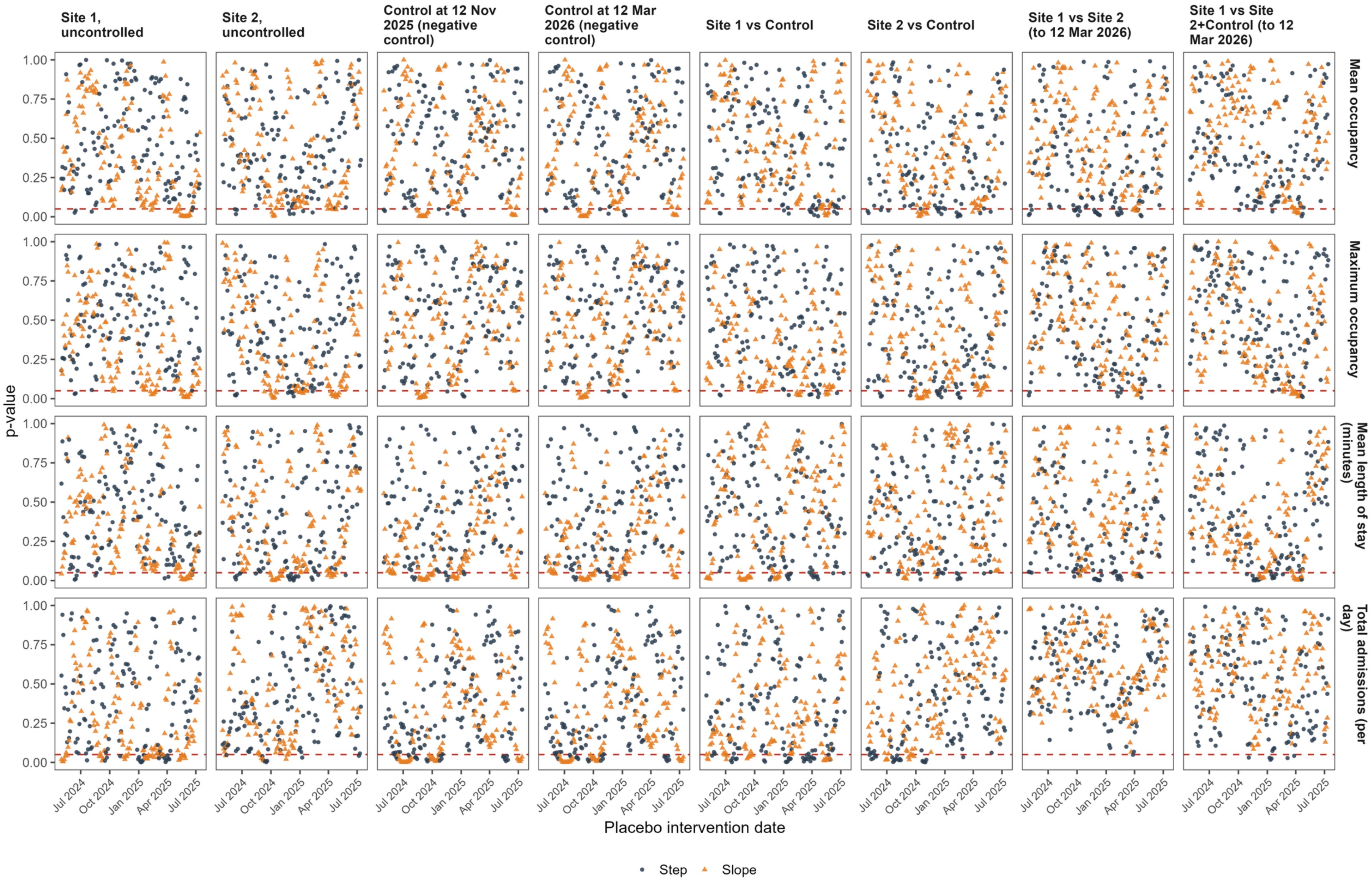
